# Epidemiological impact of SARS-CoV-2 vaccination: mathematical modeling analyses

**DOI:** 10.1101/2020.04.19.20070805

**Authors:** Monia Makhoul, Houssein H. Ayoub, Hiam Chemaitelly, Shaheen Seedat, Ghina R Mumtaz, Sarah Al-Omari, Laith J. Abu-Raddad

## Abstract

**Background:** Several SARS-CoV-2 vaccine candidates are currently in the pipeline. This study aims to inform SARS-CoV-2 vaccine development, licensure, decision-making, and implementation by determining key preferred vaccine product characteristics and associated population-level impact.

**Methods:** Vaccination impact was assessed at various efficacies using an age-structured mathematical model describing SARS-CoV-2 transmission and disease progression, with application for China.

**Results:** A prophylactic vaccine with efficacy against acquisition (*VE*_*S*_) of ≥70% is needed to eliminate this infection. A vaccine with *VE*_*S*_ <70% will still have a major impact, and may control the infection if it reduces infectiousness or infection duration among those vaccinated who acquire the infection, or alternatively if supplemented with a moderate social-distancing intervention (<20% reduction in contact rate), or complemented with herd immunity. Vaccination is cost-effective. For a vaccine with *VE*_*S*_ of 50%, number of vaccinations needed to avert one infection is only 2.4, one severe disease case is 25.5, one critical disease case is 33.2, and one death is 65.1. Gains in effectiveness are achieved by initially prioritizing those ≥60 years. Probability of a major outbreak is virtually zero with a vaccine with *VE*_*S*_ ≥70%, regardless of number of virus introductions. Yet, an increase in social contact rate among those vaccinated (behavior compensation) can undermine vaccine impact.

**Conclusions:** Even a partially-efficacious vaccine can offer a fundamental solution to control SARS-CoV-2 infection and at high cost-effectiveness. In addition to the primary endpoint on infection acquisition, developers should assess natural history and disease progression outcomes and/or proxy biomarkers, since such secondary endpoints may prove critical in licensure, decision-making, and vaccine impact.

## Introduction

Following the Severe Acute Respiratory Syndrome (SARS) epidemic in 2002 and the Middle East Respiratory Syndrome (MERS) epidemic in 2012 [1], a novel coronavirus, severe acute respiratory syndrome coronavirus 2 (SARS-CoV-2), emerged in late December 2019 in Wuhan, Hubei province, China [2, 3]. While the earlier coronavirus epidemics were rather limited in scope and scale [1], SARS-CoV-2 rapidly spread [4] and evolved into a pandemic [5].

In absence of a vaccine, even if partially efficacious [6], containment of the epidemic in China necessitated large-scale contact tracing and testing through deployment of thousands of healthcare fieldworkers along with severe quarantine measures [4]. The strain put on healthcare systems [7], and the global human [8, 9] and economic [10] losses caused by the virus and resulting disease, designated as Coronavirus Disease 2019 (COVID-2019) [11], accelerated efforts towards vaccine development [6, 12]. While multiple vaccine candidates are currently in the pipeline, they are still in early stage of development [6, 12, 13].

Assessment of the population-level impact of vaccine candidates through mathematical modeling is a critical component of the process of vaccine development, value proposition, licensure, decision-making, and pathways and costs of vaccine administration, and has been utilized for a wide range of infectious diseases [14-28]. In early stages of development, modeling is used to define the vaccine’s key preferred product characteristics, by estimating levels of efficacy necessary to observe significant population-level impact, determining necessary duration of protection/immunity incurred by the vaccine, and identifying priority populations for optimal effectiveness [21, 29, 30]. These parameters provide early guidance to developers, manufacturers, regulators, and decision makers about candidates that are likely to be optimal through specifying vaccine characteristics that will maximize public health impact and cost-effectiveness [21, 28, 29, 31, 32]. Once key attributes are established, modeling plays an integral role in building the case for investment in vaccine development, and in ensuring rapid roll-out post-licensing, through assessment of risks, costs, and predicted returns associated with different immunization strategies [29, 33]. Post-vaccination, modeling is used to inform design and interpretation of surveillance studies [25-27].

We aimed in this study to provide the scientific evidence necessary to inform and accelerate SARS-CoV-2 vaccine development, licensure, decision-making, and implementation by determining key preferred vaccine product characteristics and associated population-level impact, at a critical time for such development [6, 12, 13].

## Methods

### Mathematical Model

A deterministic model was constructed to describe SARS-CoV-2 transmission dynamics in a given population, here China as an illustrative example, in presence of vaccination (Figure S1 of Supplementary Material (SM)). The model extended a recently-developed age-structured model focused on analyzing SARS-CoV-2 epidemiology in China [34], the country with the most complete epidemic cycle to date. Model structure was informed by current understanding of SARS-CoV-2 natural history and epidemiology, and consisted of a set of coupled nonlinear differential equations that stratify the population into compartments based on vaccination status, age group, infection status, infection stage, and disease stage (Section A of SM).

For each of vaccinated and unvaccinated populations, nine age groups were considered, each representing a 10-year age band except for the last category (0-9, 10-19, …, ≥80 years). Susceptible individuals were at risk of being exposed to the infection at varying hazard rates depending on their age group and vaccination status. Following a latency period, infected individuals develop asymptomatic or mild infection followed by recovery, or severe infection followed by severe disease and then recovery, or critical infection followed by critical disease and either recovery or disease mortality. Mixing between individuals of different age groups was determined by an age mixing matrix that allows a range of mixing. Details on model structure are in SM. The model was coded, fitted, and analysed using MATLAB R2019a [35].

### Model parametrization and fitting

The model was parameterized and calibrated using empirical data on SARS-CoV-2 natural history and epidemiology. Age-specific distributions of infected individuals across the mild, severe, or critical infection stages were based on case-severity levels observed in China [4, 36, 37]. Critical disease cases were at risk of disease mortality, with the relative mortality rate in each age group informed by the age-specific crude case fatality rate observed in China [3, 38]. Population size, demographic structure (age distribution), and life expectancy, as of 2020, were obtained from the United Nations World Population Prospects database [39]. The model was fitted to empirical time series data for the daily and cumulative numbers of diagnosed SARS-CoV-2 cases and deaths [40], number of recovered individuals [40], as well as the age-specific attack rate [38, 41]. Details of model parameters, values, and justifications are in Tables S1 and S2 and Section B of SM.

### Product characteristics of candidate vaccines

We assessed the impact of a prophylactic vaccine that reduces susceptibility to infection. However, since the first available vaccine may only be partially efficacious against infection acquisition, we also assessed the impact of the vaccine assuming additional “breakthrough” effects, that is effects that modulate natural history of infection for those vaccinated but still acquire the infection. Specifically, we assumed that vaccination may reduce infectiousness per one contact (by reducing viral load), infection duration (by faster clearance with vaccine-induced immunity), and likelihood of developing severe or critical disease (by rapid immune response that prevents disease progression). Definitions of these vaccine efficacies are summarized in Table 1.

**Table 1.** Key vaccine product characteristics used to assess impact of a vaccine against SARS-CoV-2.

| **Vaccine characteristic** | **Definition** | **Description** |
| --- | --- | --- |
| $VE_s$ | Vaccine efficacy in reducing susceptibility | Proportional reduction in the susceptibility to infection acquisition among those vaccinated compared to those unvaccinated |
| $VE_I$ | Vaccine efficacy in reducing infectiousness | Proportional reduction in infectiousness (lower viral load due to vaccine-primed immune response) among those vaccinated but still acquired the infection compared to those unvaccinated |
| $VE_{P_1}$ | Vaccine efficacy in reducing duration of infection | Proportional reduction in the duration of mild infection (faster infection clearance due to vaccine-primed immune response) among those vaccinated but still acquired the infection compared to those unvaccinated |
| $VE_{P_2}$ | Vaccine efficacy in reducing the fraction of individuals with severe or critical infection | Proportional reduction in the fraction of individuals with severe or critical infection (lower probability of developing severe or critical infection due to vaccine-primed immune response) among those vaccinated but still acquired the infection compared to those unvaccinated |
| $D$ | Duration of vaccine protection | Duration of protection that the vaccine will elicit |
| $r$ | Behavior compensation post-vaccination | Proportional increase in social contact rate (reduced social distancing) among those vaccinated compared to those unvaccinated |

Other relevant characteristics include duration of protection elicited by the vaccine and vaccination effect on adherence to social distancing—we investigated impact of increasing social contact rate following vaccination with the perception of protection.

### Measures of vaccine impact

Population-level impact of SARS-CoV-2 vaccination was assessed by quantifying incidence, cumulative incidence, and reduction in incidence of each of infections, severe disease cases, critical disease cases, and deaths arising in presence of vaccination compared to the counter-factual scenario of no-vaccination. Vaccination impact was further assessed by quantifying *effectiveness*, that is number of vaccinations needed to avert one infection or one adverse disease outcome (ratio of number of vaccinations relative to that of averted outcomes). The latter measure is essentially cost-effectiveness with no costs included, as they are not yet available. Vaccination impact was assessed at: 1) *VE_S_* = 50% but 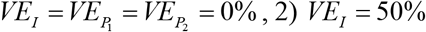 but 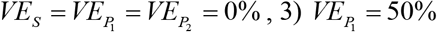 but 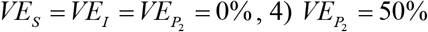 but 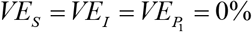 and 5) 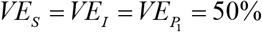but. Vaccine was assumed to elicit protection over 10 years.

### Vaccination program scenarios

Two vaccination program scenarios were considered. In both programs, it was assumed that vaccination is introduced in absence of social-distancing interventions, as the purpose of vaccination is to replace such interventions. The first program scenario assumes vaccine introduction and scale-up to 80% coverage before epidemic onset. This scenario is relevant for assessing impact of vaccination on future SARS-CoV-2 introductions in countries where the epidemic has been contained or at low level, such as in China. The scenario is also relevant to assess *maximum* potential impact of vaccination regardless of current epidemic status. The second program scenario assumes vaccine introduction during the epidemic’s exponential growth phase, with scale-up to 80% coverage within one month.

### Additional analyses

Incidence of new infections was assessed at various levels of *VE*_*S*_ to determine the *minimum* efficacy needed to fully control the infection. Incidence was also assessed in a scenario where vaccination was introduced with a social-distancing intervention to estimate level of social distancing needed to complement vaccination to control the infection. Incidence was assessed in another scenario where those vaccinated increased their social contacts (behavior compensation), to assess consequences on vaccination impact. Lastly, we derived and estimated likelihood of occurrence of a major outbreak following infection introduction in a vaccinated but infection-free population (Section D of SM).

### Uncertainty analysis

A multivariable uncertainty analysis was conducted to determine range of uncertainty around model predictions using five-hundred model runs. At each run, Latin Hypercube sampling [42, 43] was applied in selecting the natural history and disease progression parameter values from ranges specified by assuming ±30% uncertainty around parameters’ point estimates. The model was then refitted to input data and vaccine impact assessed in the new fitted model. Resulting distribution for vaccine impact across all 500 runs was used to calculate predicted means and 95% uncertainty intervals (UIs).

## Results

Figures 1 and 2 illustrate impact of vaccination assuming different vaccine product characteristics (efficacies) for each vaccination program roll-out scenario. In the first scenario (Figure 1 and Figures S2 and S3 of SM), where vaccination was scaled up to 80% coverage before epidemic onset, the epidemic in absence of vaccination peaked at 158 days after virus introduction, but at 286 days when 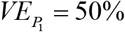, 452 days when *VE*_*I*_ = 50%, and 462 days when *VE*_*S*_ = 50%. There was no epidemic when 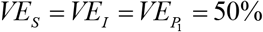. A vaccine with *VE*_*S*_ = 50% reduced peak infection incidence by 84.4% and cumulative/total infections by 52.8%, peak severe disease incidence by 83.9% and cumulative severe disease cases by 53.4%, peak critical disease incidence by 82.1% and cumulative critical disease cases by 46.7%, peak death incidence by 79.0% and cumulative deaths by 44.4%. A vaccine with *VE*_*I*_ = 50% yielded slightly lower reductions in incidence of infection and adverse outcomes, while a vaccine with 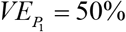 was less impactful though still achieved considerable reductions. A vaccine with 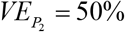 had no impact on infection incidence, but reduced peak incidence of each of severe and critical disease by 38.5%, and deaths by 40.0% (Figure 1), and the cumulative incidence of the latter three outcomes by ∼39% (Figures S2 and S3 of SM).

**Figure 1.**
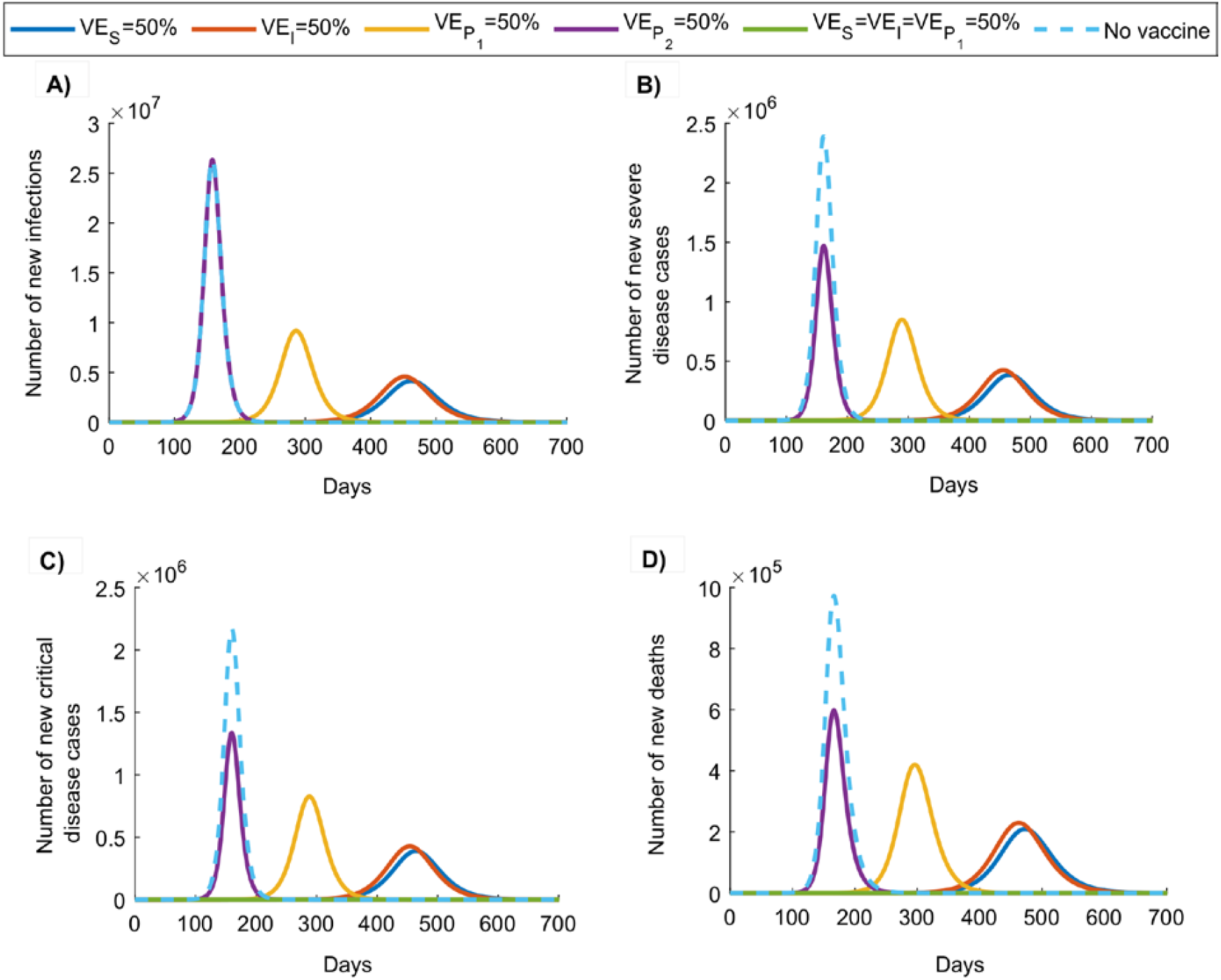
Impact of SARS-CoV-2 vaccination on number of A) new infections, B) new severe disease cases, C) new critical disease cases, and D) new deaths in the scenario assuming vaccine scale-up to 80% coverage before epidemic onset. Duration of vaccine protection is 10 years. Impact was assessed at 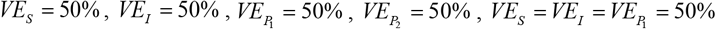.

**Figure 2.**
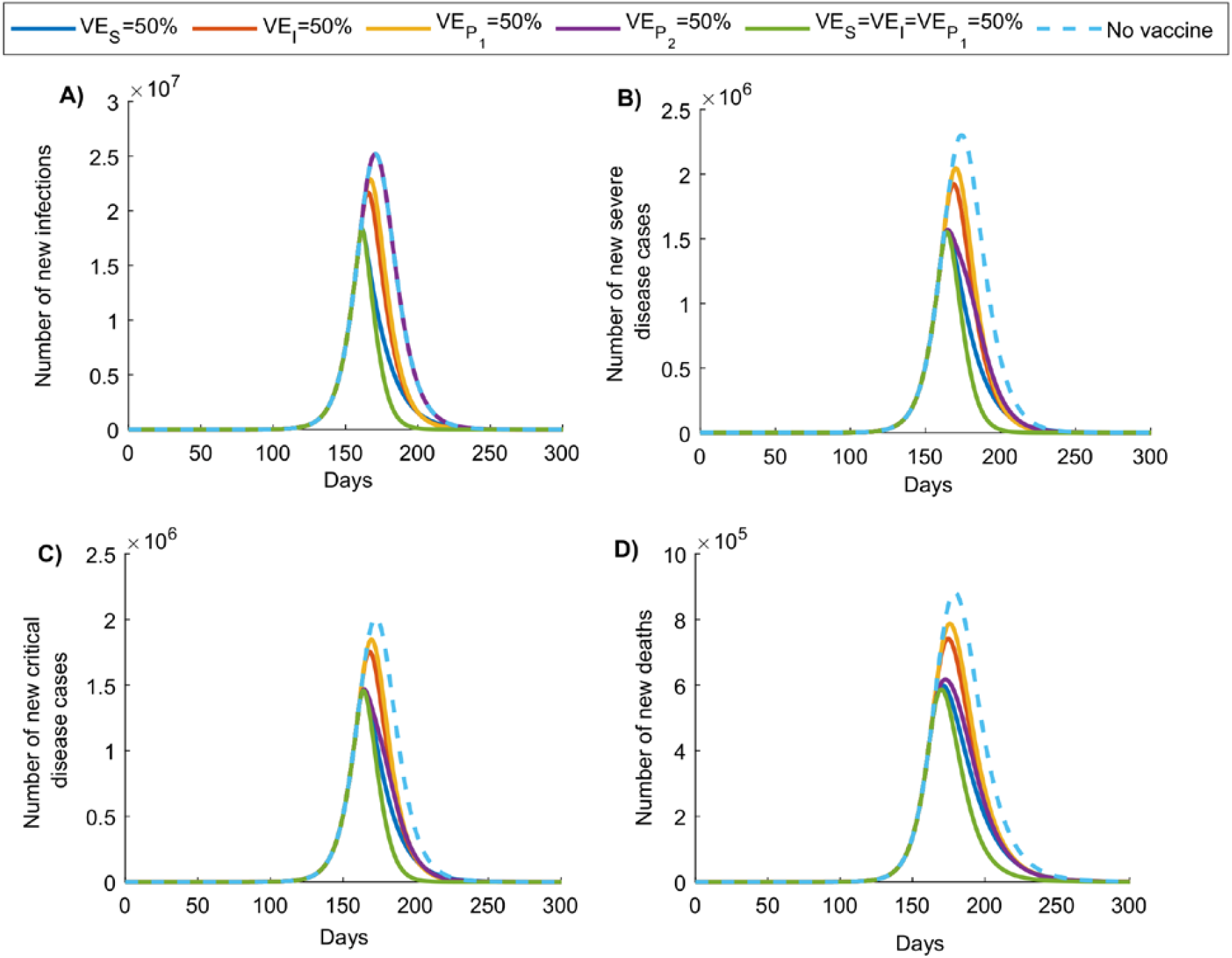
Impact of SARS-CoV-2 vaccination on number of A) new infections, B) new severe disease cases, C) new critical disease cases, and D) new deaths in the scenario assuming vaccine introduction during the exponential growth phase of the epidemic, with scale-up to 80% coverage within one month. Duration of vaccine protection is 10 years. Impact was assessed at 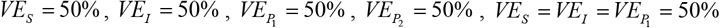.

In the second scenario (Figure 2 and Figures S4 and S5 of SM), where vaccination was rapidly scaled up to 80% coverage during the exponential growth phase, the epidemic peaked earlier and at lower values for incidence of infection and adverse outcomes. Impact of a vaccine with *VE*_*S*_ = 50% was initially similar to that of a vaccine with 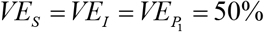, however, over time, the latter was more impactful in reducing infection and adverse outcomes. Reduction in cumulative number of new infections (at end of epidemic cycle) was highest for 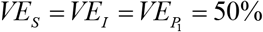 at 53.4%, followed by *VE*_*S*_ = 50% at 41.2%, *VE*_*I*_ = 50% at 28.2%, and 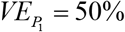 at 23.1%, with no reduction for 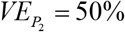. Reduction in cumulative number of new deaths for these efficacies was, respectively, 47.2%, 34.8%, 22.5%, 18.0%, and 30.0%. Figure 3 illustrates vaccine effectiveness in averting infection and adverse outcomes by end of epidemic cycle for first program scenario. For *VE*_*S*_ = 50%, 2.4 vaccinations were needed to avert one infection, 25.4 to avert one severe disease case, 33.2 to avert one critical disease case, and 65.1 to avert one death. Effectiveness was nearly comparable for *VE*_*I*_ = 50%, whereas more vaccinations were needed to avert one infection or one adverse outcome for 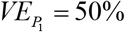 and 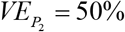. Best effectiveness was for 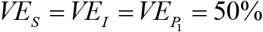 where only 1.3 vaccinations were needed to avert one infection, 13.6 to avert one severe disease case, 15.5 to avert one critical disease case, and 28.9 to avert one death. Graphs illustrating temporal evolution of vaccine effectiveness for vaccination program scenarios 1 and 2 are in Figures S6 and S7 of SM, respectively.

**Figure 3.**
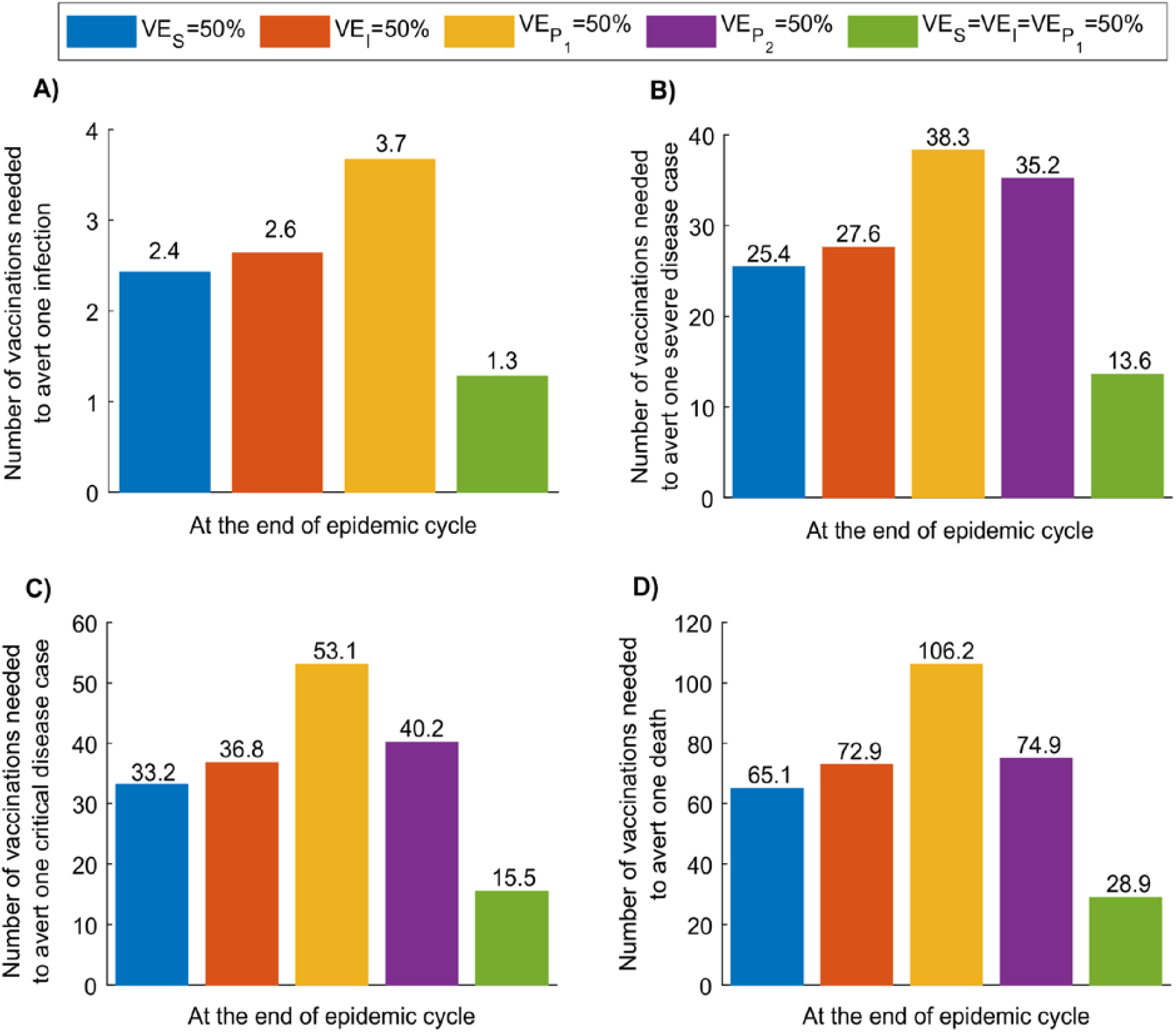
SARS-CoV-2 vaccine effectiveness. Number of vaccinations needed to avert A) one infection, B) one severe disease case, C) one critical disease case, and D) one death, by end of epidemic cycle. Scenario assumes vaccine scale-up to 80% coverage before epidemic onset. Duration of vaccine protection is 10 years. Impact was assessed at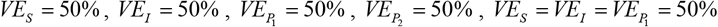. Panel A does not include the result for 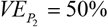, as this efficacy has no impact on number of infections—it affects only severe and critical disease and death.

Figure 4 shows effectiveness of age-group prioritization by end of epidemic cycle using a vaccine with *VE*_*S*_ = 50%, introduced before epidemic onset. Prioritizing adults ≥20 years of age was most effective in reducing infection incidence, with ≤3 vaccinations needed to avert one infection. Meanwhile, prioritizing adults ≥60 years of age was most effective in reducing new deaths, with ≤36 vaccinations needed to avert one death. Prioritizing children was least effective with large number of vaccinations needed to avert one infection or one adverse outcome. Of note that there are minor differences in effectiveness over time. For instance, in initial phases of the epidemic, prioritizing those 60-69 years of age was slightly more effective than prioritizing those 40-49 years of age (Figure S8 of SM). Meanwhile, towards end of epidemic cycle, the inverse was true.

**Figure 4.**
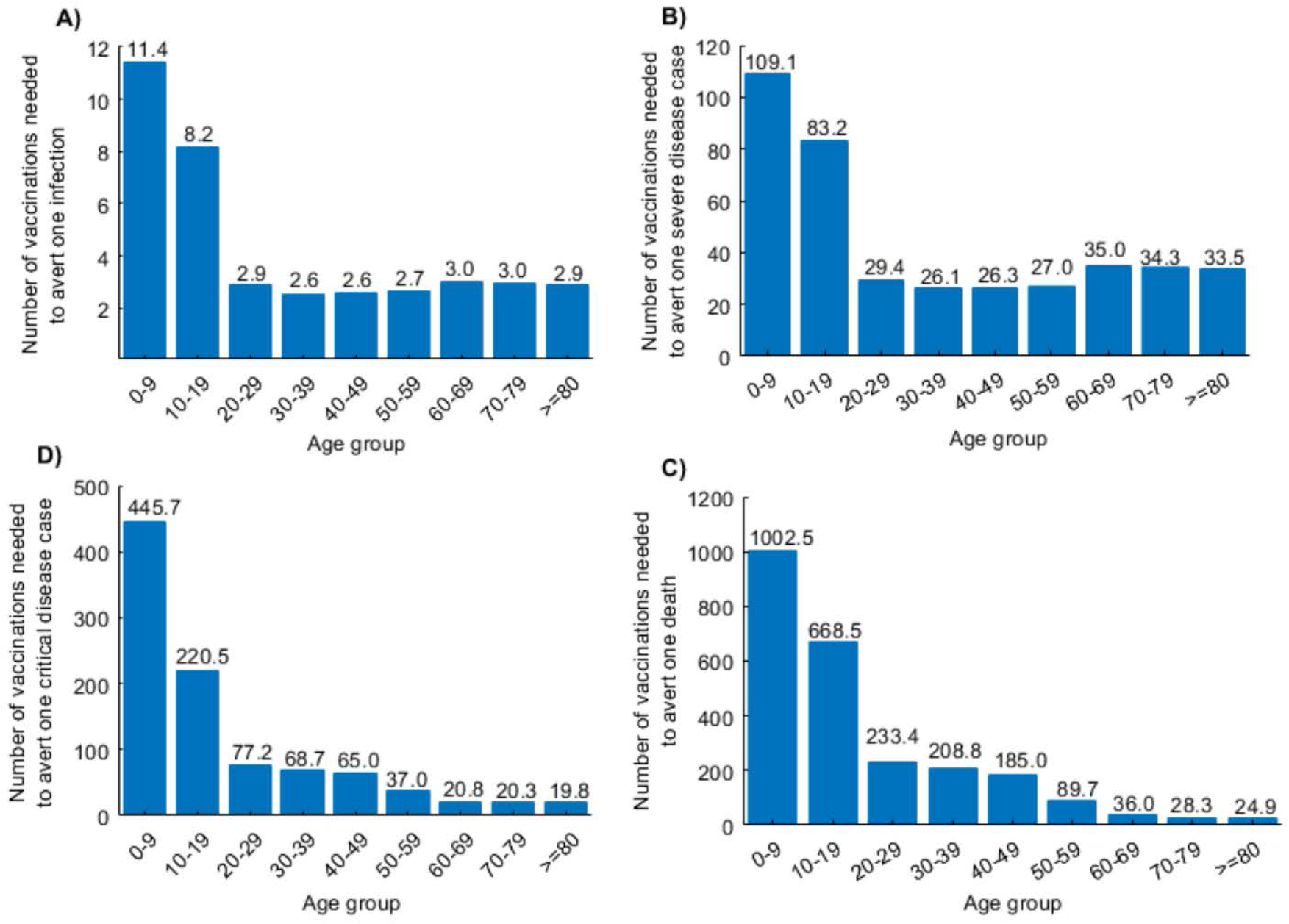
Effectiveness of age-group prioritization using a SARS-CoV-2 vaccine with *VE*_*s*_ of 50%. Number of vaccinations needed to avert A) one infection, B) one severe disease case, C) one critical disease case, and D) one death, by prioritizing different age groups for vaccination. Scenario assumes vaccine scale-up to 80% coverage before epidemic onset, and duration of vaccine protection of 10 years. Effectiveness is assessed at end of epidemic cycle.

Figure 5A shows cumulative number of infections (at end of epidemic cycle) at various *VE*_*S*_ levels for a vaccine introduced before epidemic onset. A gradual decrease is observed as *VE*_*S*_ increases, with an accelerated reduction as *VE*_*S*_ approaches 60%—the level beyond which number of infections approaches zero. Epidemic onset is prevented at *VE*_*S*_ = 69%. Figure 5B illustrates the gains in effectiveness as *VE*_*S*_ increases, with 5.3 vaccinations needed to avert one infection at *VE*_*S*_ = 30%, but only 1.3 at *VE*_*S*_ = 60%.

**Figure 5.**
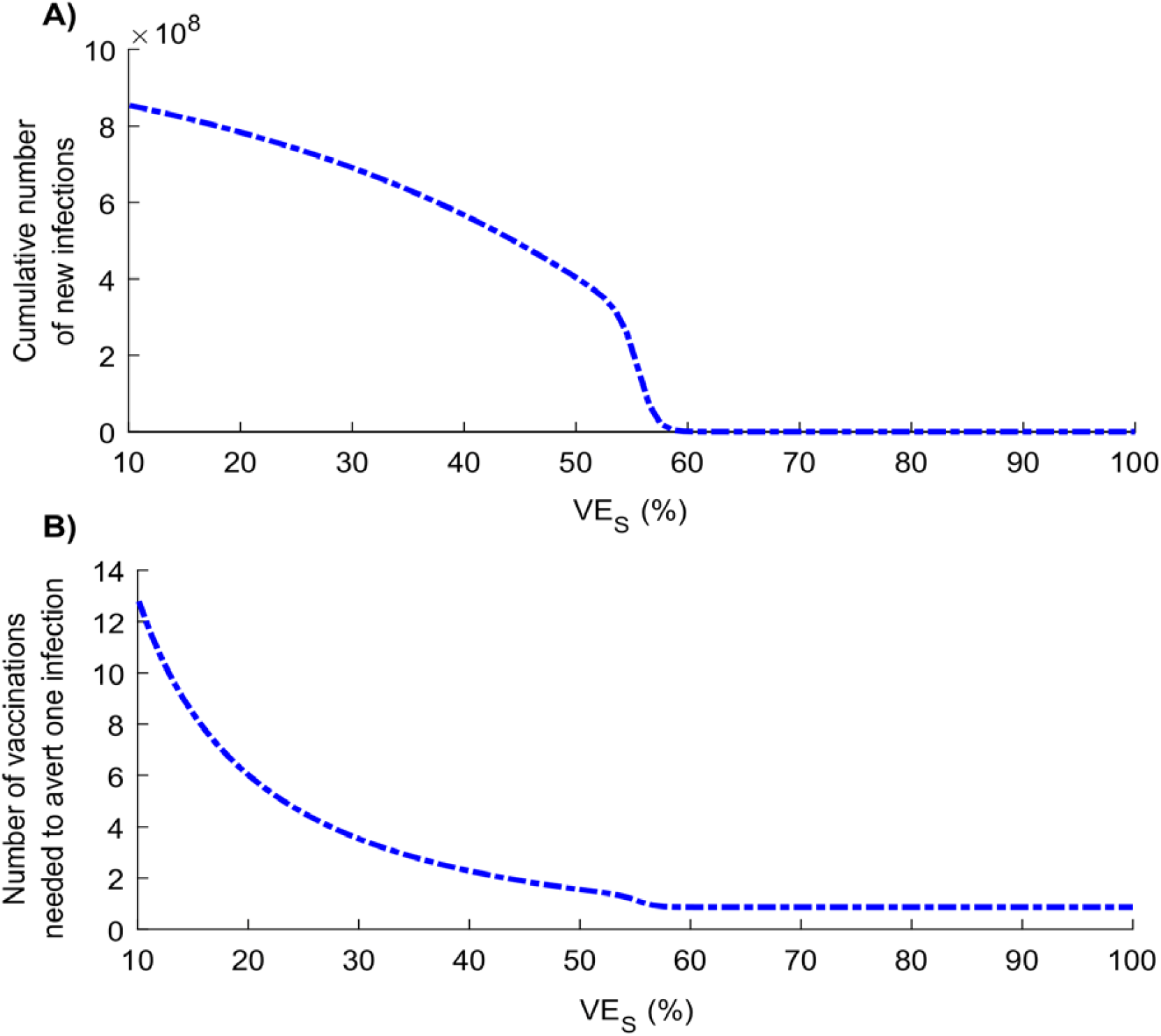
Impact of varying levels of vaccine efficacy in reducing susceptibility (*VE*_*s*_) on A) cumulative number of new SARS-CoV-2 infections and B) number of vaccinations needed to avert one SARS-CoV-2 infection. Scenario assumes vaccine scale-up to 80% coverage before epidemic onset. Duration of vaccine protection is 10 years. Measures are assessed at end of epidemic cycle.

While a vaccine with *VE*_*S*_ = 50% cannot fully control the epidemic, Figure S9 of SM shows the impact when vaccination is supplemented with a social-distancing intervention that reduces the contact rate. Less than 20% reduction in contact rate would be sufficient to fully control the epidemic.

Vaccinated individuals may increase their contact rate with the perception of protection. Figure 6 shows the consequences of behavior compensation. A 20% increase in contact rate among those vaccinated lowered the reduction in cumulative incidence from 52.8% to only 21.0%. A 41.8% increase in contact rate nullified impact of vaccination in reducing incidence.

**Figure 6.**
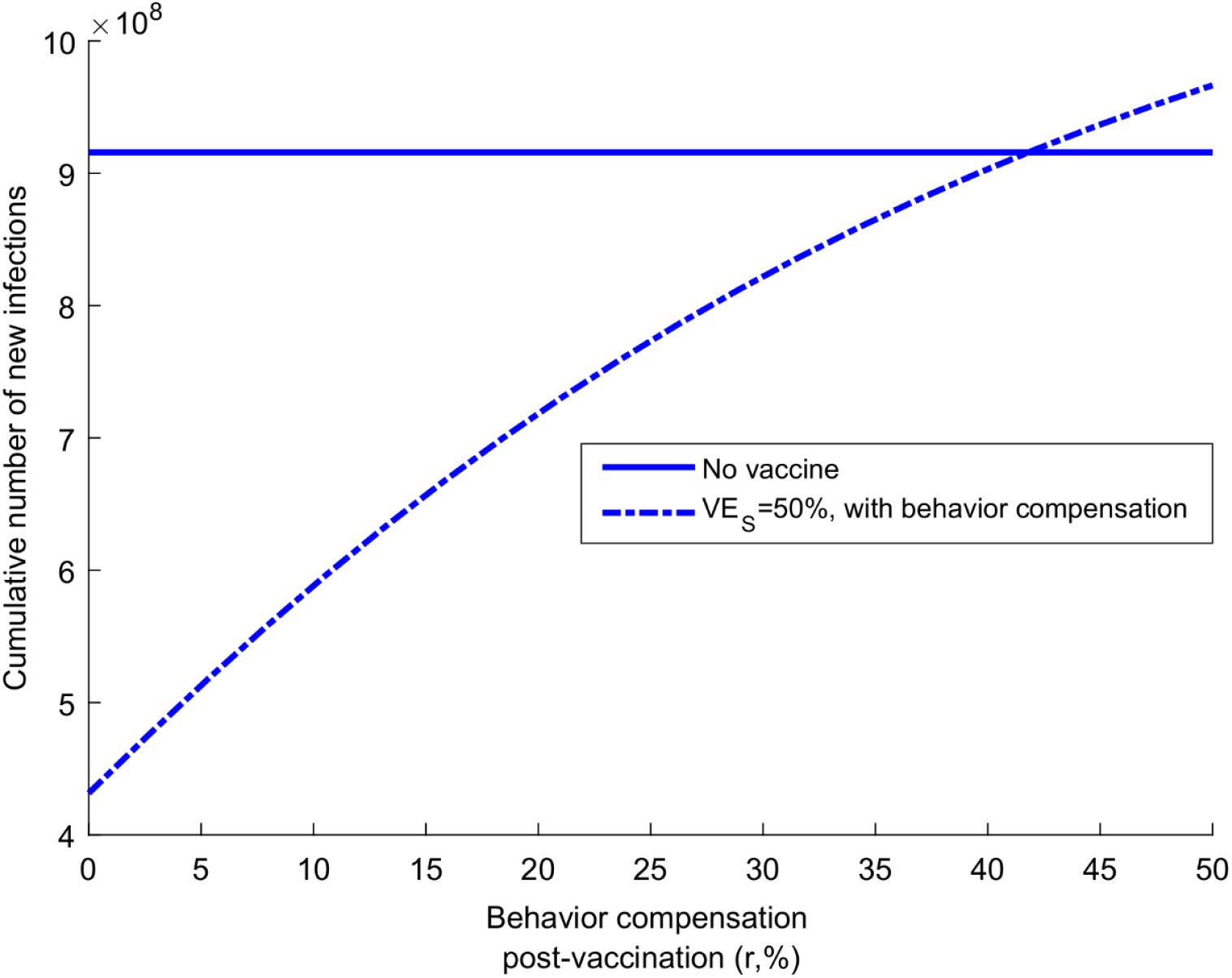
Impact of vaccination with reduced adherence to social distancing for those vaccinated. Figure shows impact of varying levels of behavior compensation post-vaccination on the vaccine-induced reduction in cumulative number of new SARS-CoV-2 infections by end of epidemic cycle. Scenario assumes vaccine scale-up to 80% coverage before epidemic onset, *VE*_*s*_ is 50%, and duration of vaccine protection is 10 years.

Figure 7 reports probability of occurrence of a major outbreak at varying levels of vaccine efficacy. A gradual decrease is noted with the increase in efficacy that accelerates close to *VE*_*S*_ or *VE*_*I*_ of 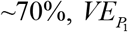 85%, and 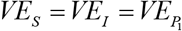 of ∼33% beyond which no major outbreak is expected to occur. Of note that this figure shows (conservatively) the upper bound of the probability of a major outbreak. Results for the lower bound are in Figure S10 of SM and derivations in Section D of SM.

**Figure 7.**
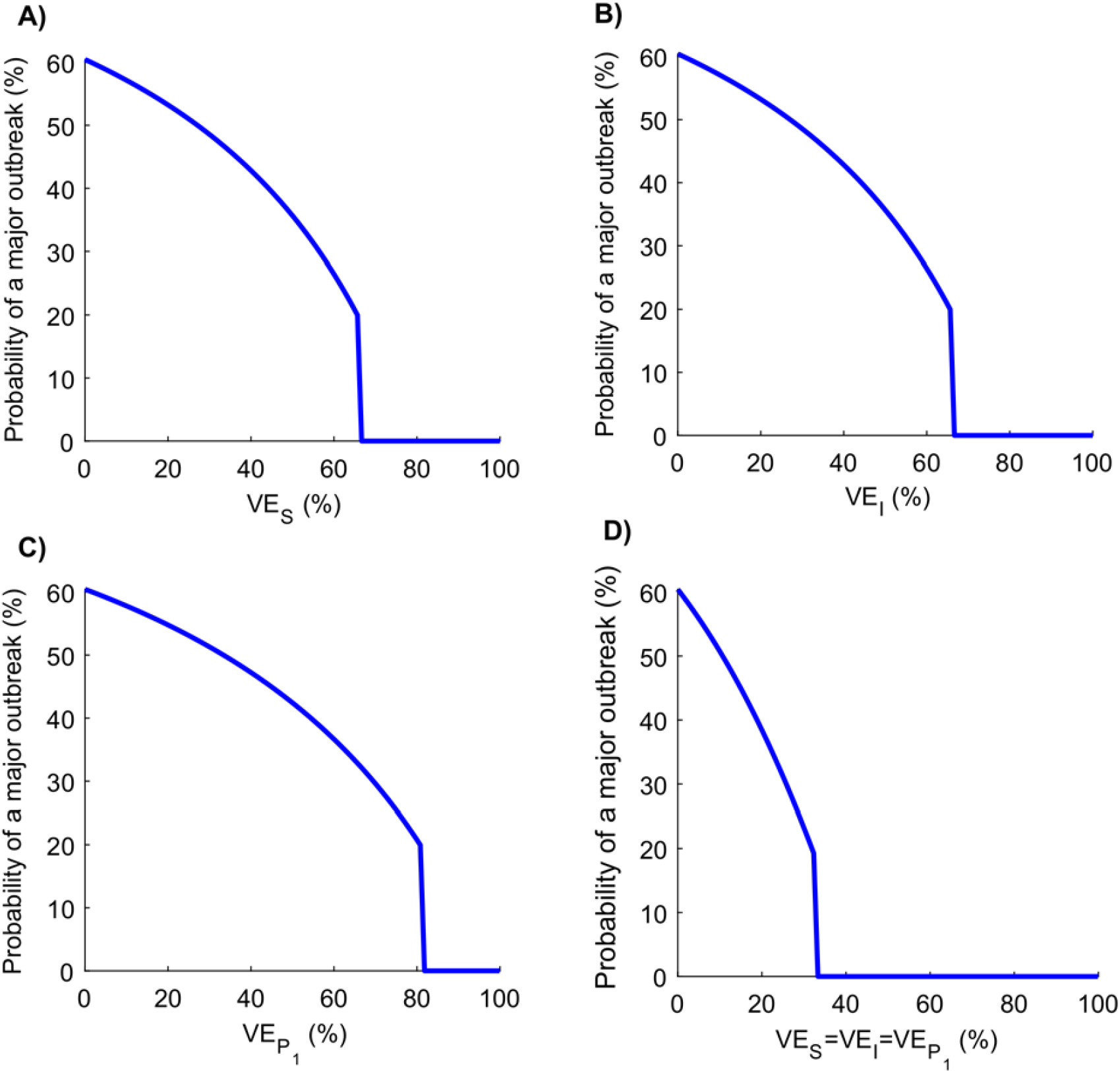
Probability of occurrence of a major outbreak following vaccination. Probability of occurrence of a major outbreak upon virus introduction at varying levels of A) *VE_S_*, B) *VE_I_*, C)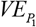, and D)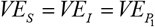. Scenario assumes vaccine scale-up to 80% coverage before epidemic onset. Duration of vaccine protection is 10 years. Figure does not include the result for 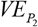, as this efficacy has no impact on probability of occurrence of a major outbreak.

Uncertainty analysis demonstrated robustness of model predictions to a wide range of uncertainty in input parameters (Figure S11 of SM).

## Discussion

Above results indicate that even a partially-efficacious vaccine can offer a fundamental solution to the SARS-CoV-2 pandemic—the vaccine does not need to have sterilizing immunity to fully control the infection. Indeed, a vaccine with *VE*_*S*_ ≥70% could be sufficient to control the pandemic at ≥80% coverage (Figure 5A). Even a vaccine with *VE*_*S*_ <70% may still control the infection if it additionally (and plausibly) induces “breakthrough” effects such as reduction in viral load (reduction in infectiousness; *VE*_*I*_) or faster infection clearance (reduction in infection duration; 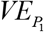) among those vaccinated who still acquire the infection. The latter effects have *individually* (that is in absence of protection against acquisition) a comparable impact on transmission to that of a prophylactic vaccine that reduces infection acquisition (Figures 1 and 2). Even in absence of such effects, infection control can still be achieved if vaccination is supplemented with only a moderate social-distancing intervention (Figure S9 of SM), or complemented with partial herd immunity—a considerable fraction of the population could have acquired the infection and developed protective antibodies by the time of vaccine roll-out. Even a vaccine that does not prevent infection yet only mitigates disease progression (reduction in severe or critical disease and death; 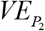), could still yield significant gains by curbing disease burden (Figures 1 and 2).

Results also indicated that vaccine impact depends on time of vaccine introduction whether before (Figure 1 and Figures S2 and S3 of SM) or after (Figure 2 and Figures S4 and S5 of SM) epidemic onset and/or growth—maximal gains are achieved with earlier introduction. Early introduction optimally defers epidemic growth, flattens the incidence curve, and reduces number of infections and disease outcomes (Figures 1 and 2 and Figures S2-S5 of SM).

The vaccine will likely be cost-effective over a broad range of efficacy levels (Figure 5B). For a vaccine with *VE*_*S*_ of 50%, number of vaccinations needed to avert one infection is only 2.4, one severe disease case is 25.5, one critical disease case is 33.2, and one death is 65.1 (Figure 3). Return on effectiveness is also rapid for such respiratory infection with fast-growing epidemic scale (Figures S6 and S7 of SM).

Effectiveness can be further enhanced by prioritizing vaccination for those ≥60 years of age, for an optimal reduction in disease cases and deaths (Figure 4). Conversely, prioritizing children is least effective, with their lower risk for developing adverse outcomes (Figure 4) [4, 44, 45]. This being said, prioritizing vaccination for any single age group, regardless of that age group, has overall lower effectiveness than extending vaccination to all age groups—vaccinating only one age group reduces the reproduction number (*R*_0_) only marginally, whereas vaccinating all age groups reduces *R*_0_ to an epidemic domain where small reductions in *R*_0_ can have more substantial impact on epidemic size (Figure S12 of SM, also Figure 3 versus Figure 4). Consequently, roll-out strategies should initially prioritize individuals ≥60 years of age, but then incrementally cover younger age cohorts, and eventually the entire population.

Vaccination will also reduce the likelihood of a major outbreak following virus introduction/reintroduction into a population (Figure 7). With a vaccine of *VE*_*S*_ ≥70%, infection transmission chains may not be sustainable, regardless of number of virus introductions. Of concern, however, is the potential increase in social contact rate among those vaccinated (behavior compensation)—a 42% increase in contact rate can virtually nullify the gains of a vaccine with *VE*_*S*_ of 50% at coverage of 80% (Figure 6). Roll-out of a vaccine with intermediate efficacy should be coupled with public health communication that stresses caution in social mixing following vaccination.

This study has limitations. Model estimations are contingent on validity and generalizability of input data. While we used available evidence for SARS-CoV-2 natural history and epidemiology, our understanding of the epidemiology is still evolving. We assessed vaccine impact using China as an illustrative example, given the advanced epidemic cycle, yet evidence suggests that many infections may have been undocumented in this country, particularly in the early epidemic phase [46]. This may affect some estimates, such as mortality, probably towards overestimation [34, 47]. While the *absolute* impact of the vaccine on disease severity and mortality may have been overestimated, the *relative* impact (reduction rate) is less likely to have been affected. Our baseline *R*_0_ for China was 2.1 [34], but *R*_0_ may vary across settings, thus affecting estimates for the minimum efficacy needed for infection elimination. For instance, for an *R*_0_ of 3, the minimum *VE*_*S*_ needed for elimination is about 90% (Figure S12 of SM). We assessed vaccine impact for one epidemic cycle, with no assessment of seasonality or future cycles. We assumed a long duration of vaccine protection (10 years), but this has limited impact on the predictions for one epidemic cycle, provided the duration of vaccine protection is greater than one year. Despite these limitations, our model was complex enough to factor the different key vaccine product characteristics, but also parsimonious enough to be tailored to the nature of available data. The model also generated results that are valid to a wide range of model assumptions.

## Conclusions

With most of the world’s population remaining susceptible to SARS-CoV-2 and the need to impose disruptive social-distancing interventions, vaccination is a reliable intervention in the long-term. Findings show that even a partially-efficacious vaccine provides a fundamental solution to the SARS-CoV-2 pandemic and at high cost-effectiveness. Vaccine impact and cost-effectiveness will not only depend on its efficacy in preventing infection, but can be enhanced if those vaccinated who still acquire the infection have reduced infectiousness, duration of infection, and disease severity. Vaccine developers should thus not only assess the primary endpoint of reduction in acquisition, but also other outcomes and/or proxy biomarkers including reductions in viral load and disease outcomes and speed of infection clearance for those vaccinated and unvaccinated. Totality of these primary and secondary endpoints may prove critical in the licensure process, decision-making, and vaccine impact once introduced into a population.

## Data Availability

All data are available within the manuscript and its supplementary material.

## Contributors

MM and HHA constructed, coded, and parameterized the mathematical model. MM conducted the analyses. HC supported model parametrization and analyses and wrote the first draft of the paper. LJA conceived and led the design of the study, construct and parameterization of the mathematical model, and drafting of the article. All authors contributed to discussion and interpretation of the results and to the writing of the manuscript. All authors have read and approved the final manuscript.

## Acknowledgements

This publication was made possible by NPRP grant number 9-040-3-008 and NPRP grant number 12S-0216-190094 from the Qatar National Research Fund (a member of Qatar Foundation). The authors are also grateful for support provided by the Biomedical Research Program and the Biostatistics, Epidemiology, and Biomathematics Research Core, both at Weill Cornell Medicine-Qatar. GM acknowledges support by UK Research and Innovation as part of the Global Challenges Research Fund, grant number ES/P010873/1. The statements made herein are solely the responsibility of the authors.

## Supplementary Material

### I. SARS-CoV-2 mathematical vaccine model

#### A. Model structure

We extended a recently-developed age-structured deterministic compartmental model to describe the impact of vaccination on severe acute respiratory syndrome coronavirus 2 (SARS-CoV-2) transmission dynamics and progression of the resulting disease, Coronavirus Disease 2019 (COVID-2019), in a given population. The model stratifies the unvaccinated and vaccinated populations into compartments according to age group (0-9, 10-19, 20-29,…, ≥80 years), infection status (uninfected, infected), infection stage (mild, severe, critical), and disease stage (severe, critical). Transmission and disease progression dynamics in vaccinated and unvaccinated cohorts are described using age-specific sets of nonlinear ordinary differential equations, where each age group *a* (*a* = 1, 2,…9) refers to a 10-year age band (0-9,10-19,..70-79) apart from the last group including all those aged ≥ 80 years. The model is illustrated in Figure S1.

**Figure S1.**
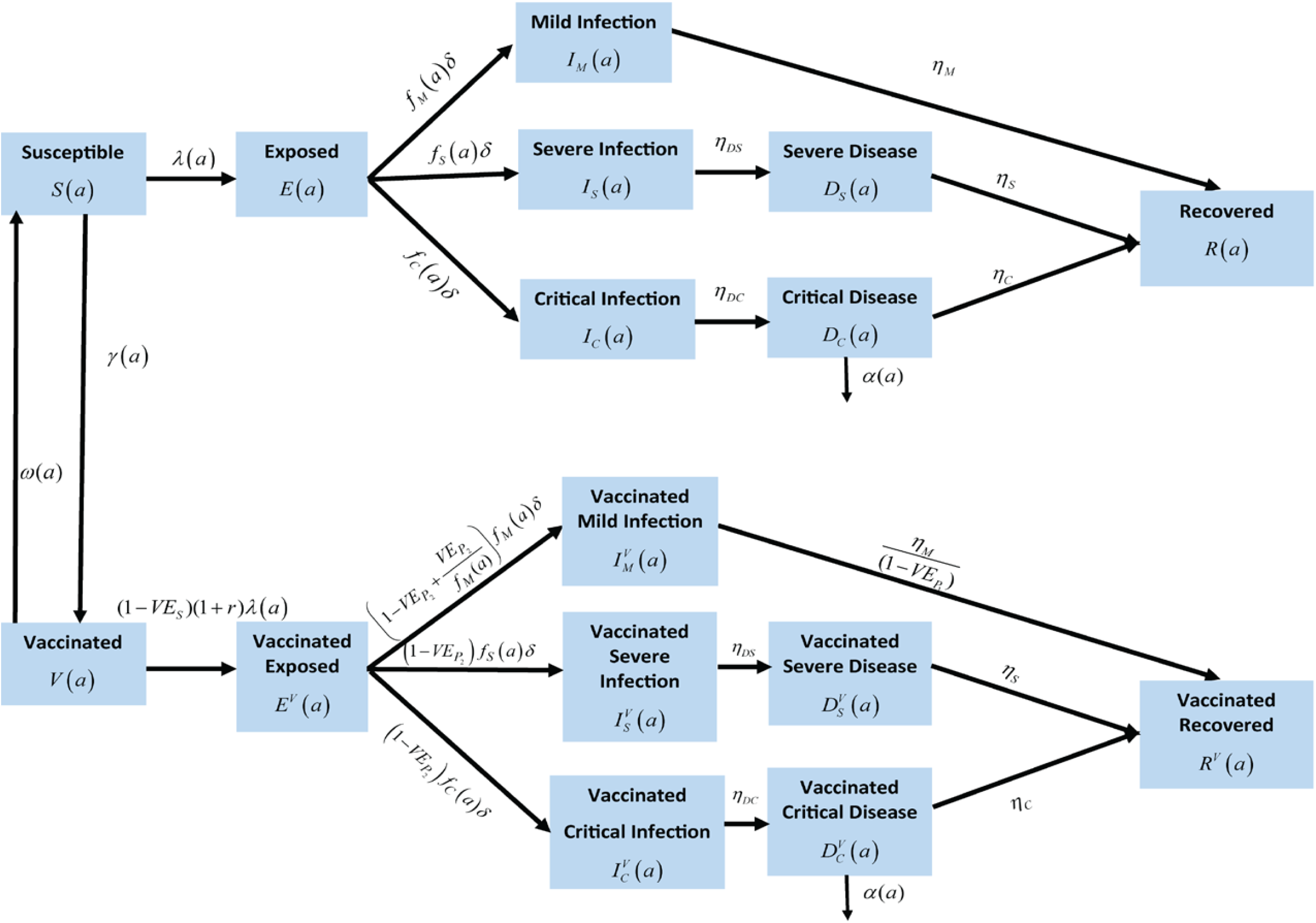
Schematic diagram describing the basic structure of the SARS-CoV-2 vaccine model.

The following set of equations describes the transmission dynamics among unvaccinated and vaccinated populations in the first age group:

#### Unvaccinated population aged 0-9 years

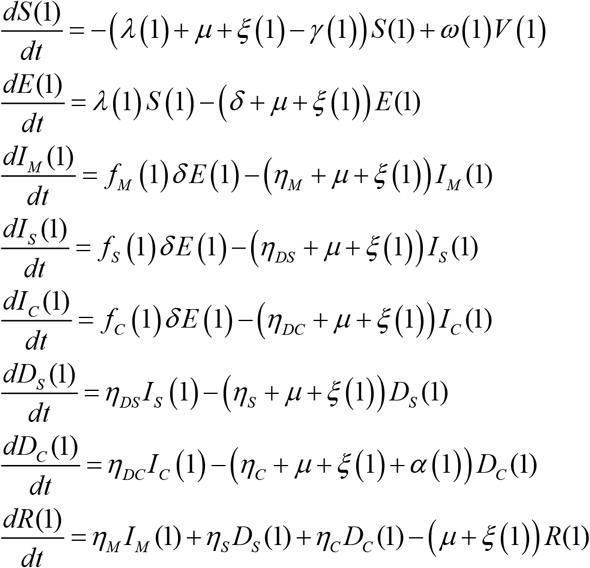

#### Vaccinated population aged 0-9 years

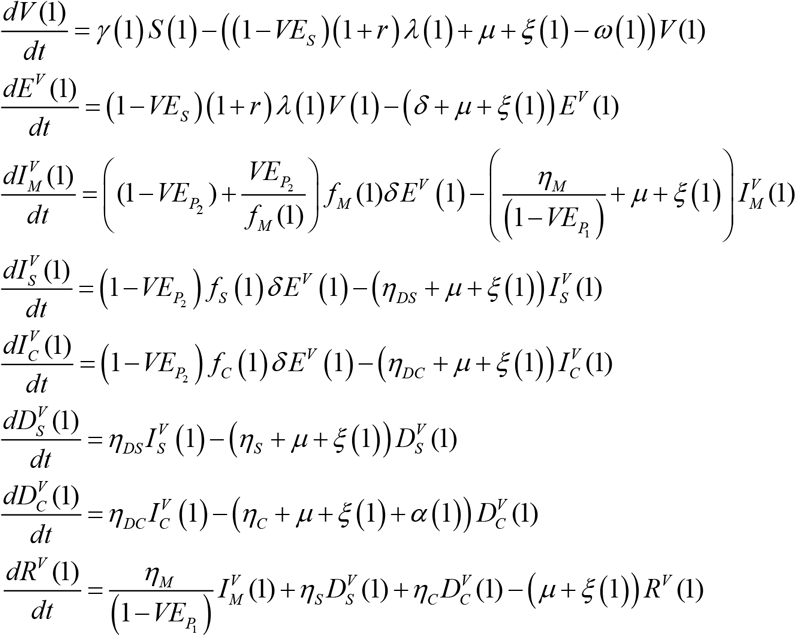

For subsequent age groups, the following set of equations was used:

#### Unvaccinated populations aged 10+ years

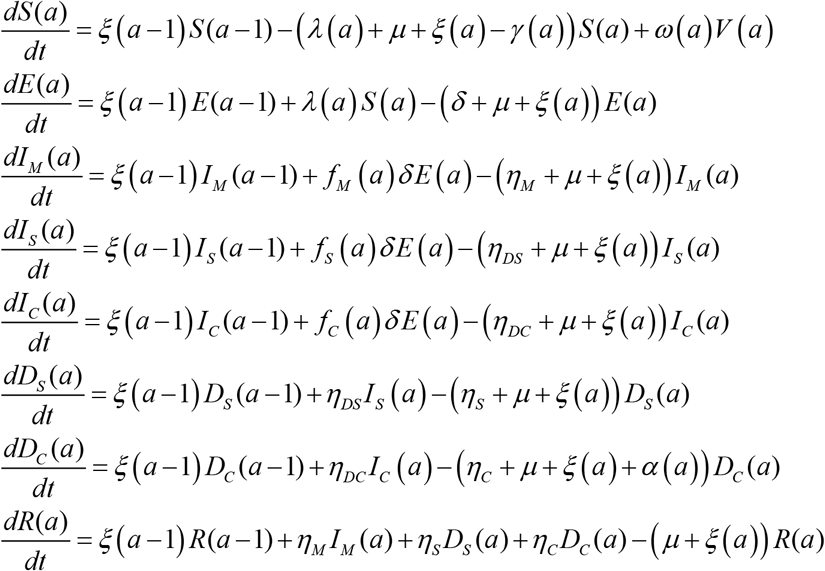

#### Vaccinated populations aged 10+ years

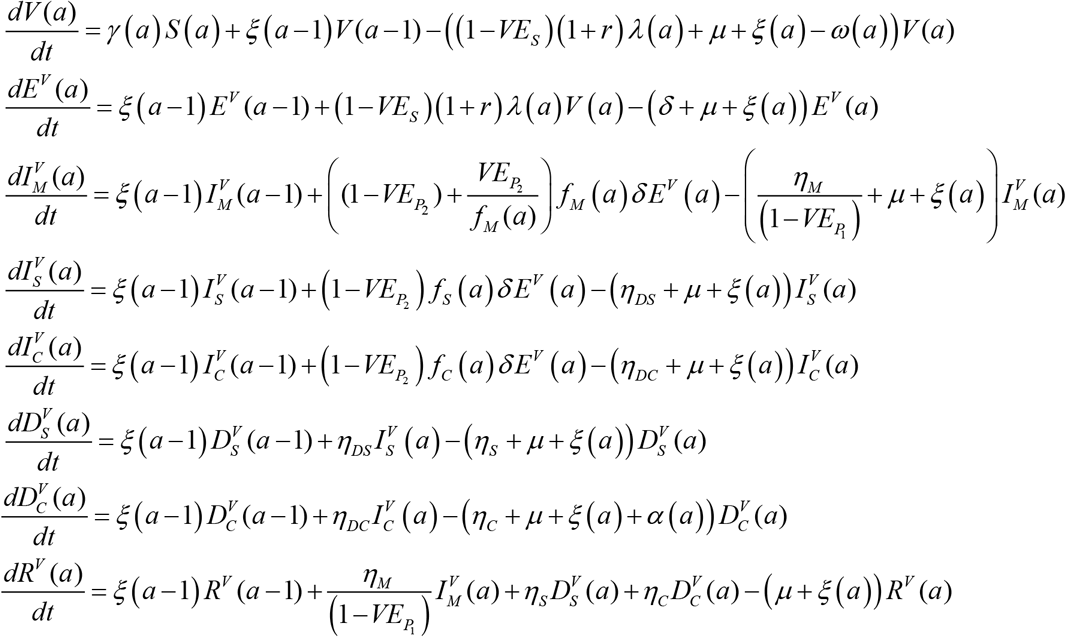

The definitions of population variables and symbols used in the equations are in Table S1.

**Table S1.** Definitions of population variables and symbols used in the model.

| Symbol | Definition |
| --- | --- |
| **Transmission dynamics parameters** |  |
| $S(a)$ | Unvaccinated susceptible population |
| $E(a)$ | Unvaccinated latently infected population |
| $I_M(a)$ | Unvaccinated population with asymptomatic or mild infection |
| $I_S(a)$ | Unvaccinated population with severe infection |
| $I_C(a)$ | Unvaccinated population with critical infection |
| $I(a)$ | Unvaccinated infected population |
| $D_S(a)$ | Unvaccinated population with severe disease |
| $D_C(a)$ | Unvaccinated population with critical disease |
| $R(a)$ | Unvaccinated recovered population |
| $V(a)$ | Vaccinated susceptible population |
| $E^V(a)$ | Vaccinated latently infected population |
| $I_M^V(a)$ | Vaccinated population with asymptomatic or mild infection |
| $I_S^V(a)$ | Vaccinated population with severe infection |
| $I_C^V(a)$ | Vaccinated population with critical infection |
| $I^V(a)$ | Vaccinated infected population |
| $D_S^V(a)$ | Vaccinated population with severe disease |
| $D_C^V(a)$ | Vaccinated population with critical disease |
| $R^V(a)$ | Vaccinated recovered population |
| $N$ | Total population size |
| $n_{age}$ | Number of age groups |
| $\xi(a)$ | Transition rate from one age group to the next age group |
| $\sigma(a)$ | Susceptibility profile to the infection in each age group |
| $\beta$ | Overall infectious contact rate |
| $1/\delta$ | Duration of latent infection |
| $1/\eta_M$ | Duration of asymptomatic or mild infection |
| $1/\eta_{DS}$ | Duration of severe infection infectiousness before isolation and/or hospitalization |
| $1/\eta_S$ | Duration of severe disease following onset of severe disease |
| $1/\eta_{DC}$ | Duration of critical infection infectiousness before isolation and/or hospitalization |
| $1/\eta_C$ | Duration of critical disease following onset of critical disease |
| $1/\mu$ | Natural death rate |
| $CFR(a)$ | *Relative* case fatality rate in each age group |
| $\alpha(a)$ | Mortality rate in each age group |
| $f_M(a)$ | Proportion of infections that will progress to be mild or asymptomatic infections |
| $f_S(a)$ | Proportion of infections that will progress to be severe infections |
| $f_C(a)$ | Proportion of infections that will progress to be critical infections |
| **Key vaccine product characteristics** |  |
| $VE_s$ | Vaccine efficacy in reducing susceptibility |
| $VE_I$ | Vaccine efficacy in reducing infectiousness |
| $VE_{P1}$ | Vaccine efficacy in reducing the duration of infection |
| $VE_{P2}$ | Vaccine efficacy in reducing the fraction of individuals with severe or critical infection |
| $1/\omega$ | Duration of vaccine protection |
| $r$ | Behavior compensation post-vaccination |

The force of infection (hazard rate of infection) experienced by the unvaccinated susceptible populations *S* (*a*) is given by

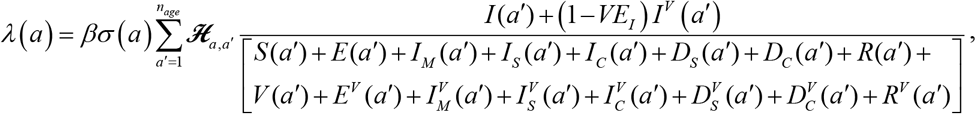

while that of vaccinated susceptible populations *V* (*a*) is given by

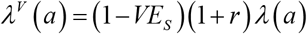

where *β* is the overall infectious contact rate. The mixing among the different age groups is dictated by the mixing matrix ℋ_*a,a*”_. This matrix provides the probability that an individual in the *a* age group will mix with an individual in the *a*′ age group (regardless of vaccination status).

The mixing matrix is given by

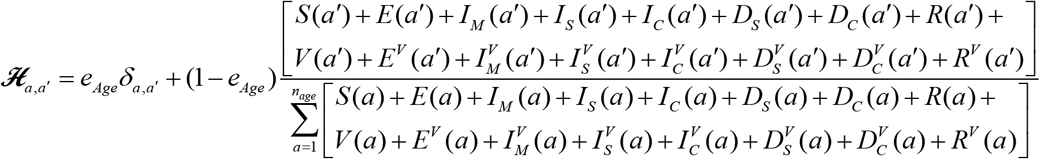

Here, *δ*_*a,a*′_ is the identity matrix. *e*_*Age*_ *∈* [0,1] measures the degree of assortativeness in the mixing. At the extreme *e*_*Age*_ = 0, the mixing is fully proportional. Meanwhile, at the other extreme, *e*_*Age*_ = 1, the mixing is fully assortative, that is individuals mix only with members in their own age group.

#### B. Parameter values

The input parameters of the model were chosen based on current empirical data for SARS-CoV-2 natural history and epidemiology. The parameter values are listed in Table S2.

**Table S2.** Model assumptions in terms of parameter values.

| Parameter | Symbol | Value | Justification |
| --- | --- | --- | --- |
| Duration of latent infection | $1/\delta$ | 3.69 days | Based on existing estimate[1] and based on a median incubation period of 5.1 days[2] adjusted by observed viral load among infected persons[3] and reported transmission before onset of symptoms[4] |
| Duration of infectiousness | $1/\eta_M$ ;<br>$1/\eta_{DS}$ ;<br>$1/\eta_{DC}$ | 3.48 days | Based on existing estimate[1] and based on observed time to recovery among persons with mild infection[1, 5] and observed viral load in infected persons[3, 4] |
| Duration of severe disease following onset of severe disease | $1/\eta_s$ | 28 days | Observed duration from onset of severe disease to recovery[5] |
| Duration of hospitalization for critical infection | $1/\eta_c$ | 28 days | Observed duration from onset of critical disease to recovery[5] |
| Life expectancy in China | $1/\mu$ | 77.47 years | United Nations World Population Prospects database[6] |
| Relative case fatality rate in each age group | $CFR(a)$ | | Observed crude case fatality rate based on China data[7, 8] |
| Age 0-9 years |  | 0% |  |
| Age 10-39 years |  | 0.2% |  |
| Age 40-49 years |  | 0.4% |  |
| Age 50-59 years |  | 1.3% |  |
| Age 60-69 years |  | 3.6% |  |
| Age 70-79 years |  | 8.0% |  |
| Age 80+ years |  | 21.9% |  |
| Proportion of infections that will progress to be mild or asymptomatic infections | $f_M(a)$ | | Observed proportion of infections that eventually develop mild or asymptomatic in China[5, 9, 10] |
| Age 0-9 years |  | 88.9% |  |
| Age 10-49 years |  | 88.0% |  |
| Age 50-59 years |  | 82.5% |  |
| Age 60+ years |  | 71.2% |  |
| Proportion of infections that will progress to be severe infections | $f_S(a)$ | | Observed proportion of infections that eventually develop severe disease in China[5, 9, 10] |
| Age 0-9 years |  | 11.1% |  |
| Age 10-49 years |  | 9.9% |  |
| Age 50-59 years |  | 10.3% |  |
| Age 60+ years |  | 7.8% |  |
| Proportion of infections that will progress to be critical infections | $f_C(a)$ | | Observed proportion of infections that eventually develop critical disease in China[5, 9, 10] |
| Age 0-9 years |  | 0.0% |  |
| Age 10-49 years |  | 2.2% |  |
| Age 50-59 years |  | 7.2% |  |
| Age 60+ years |  | 20.9% |  |
| Susceptibility profile to the infection in each age group | $\sigma(a)$ | | Estimates based on fitting the epidemic in China[11] |
| Age 0-9 years |  | 0.05 |  |
| Age 10-19 years |  | 0.05 |  |
| Age 20-29 years |  | 0.32 |  |
| Age 30-39 years |  | 0.53 |  |
| Age 40-49 years |  | 0.65 |  |
| Age 50-59 years |  | 0.74 |  |
| Age 60-69 years |  | 0.93 |  |
| Age 70-79 years |  | 0.88 |  |
| Age 80+ years |  | 0.83 |  |
| Overall infectious contact rate | $\beta$ | 0.59 contacts per day | Estimate based on fitting the epidemic in China[11] |

#### C. The basic reproduction number *R*_0_

Using the second generation matrix method described by Heffernan *et al*. [12], the basic reproduction number in absence of vaccination is given by

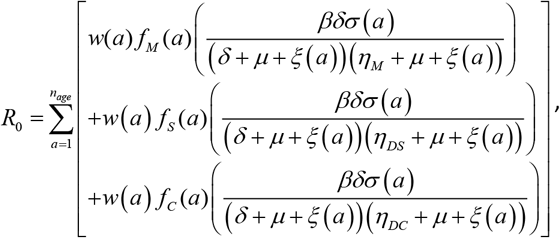

Where *w*(*a*) is the proportion of the population in each age group.

Meanwhile, the basic reproduction number in presence of vaccination is given by

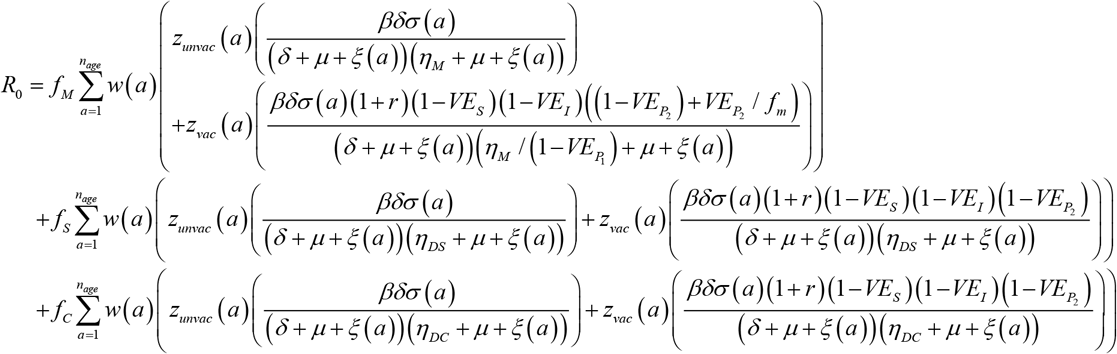

Where *z*_*unvac*_ (*a*) and *z*_*vac*_ (*a*) are, respectively, the proportions of unvaccinated and vaccinated populations in each age group.

#### D. Probability of a major outbreak

Based on Whittle’s method [13], and by constructing Bailey’s ratios [13], the probability of a major outbreak was derived, that is the probability that the fraction of susceptible individuals that become infected is ≥ *γ*, where *γ* is a specific chosen level of the final attack rate.

We defined *ρ*^*U*^_*M, N*_ (*a*), *ρ*^*U*^ _*S, N*_ (*a*), and *ρ*^*U*^ _*C, N*_ (*a*) to be, respectively, Bailey’s ratios corresponding to the unvaccinated infectious classes *I*_*M*_ (*a*), *I*_*S*_ (*a*), and *I*_*C*_ (*a*) for each age group. Similarly, we defined *ρ*^*V*^ _*M, N*_ (*a*), *ρ*^*V*^ _*S, N*_ (*a*), and *ρ*^*V*^ _*C, N*_ (*a*) to be, respectively, Bailey’s ratios corresponding to the vaccinated infectious classes 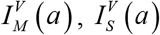, and 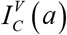 for each age group. The number of initial infections is denoted by *b*.

We define

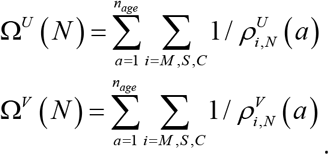

In the case of no vaccination, the probability of a major outbreak is given by the following three cases:

#### Case 1

For 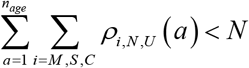, and 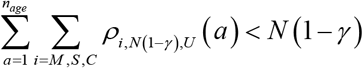, the probability of a major outbreak lies between 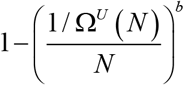 and 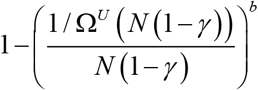

#### Case 2

For 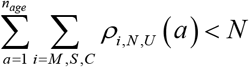, and 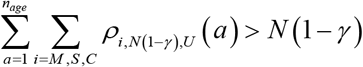, the probability of a major outbreak lies between 0 and 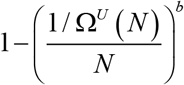.

#### Case 3

For 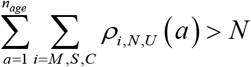, the probability of a major outbreak is 0.

Meanwhile, in the case of vaccinating a fraction of the population, the probability of a major outbreak is given by the following three cases:

#### Case 1

For 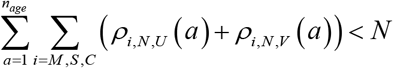 and 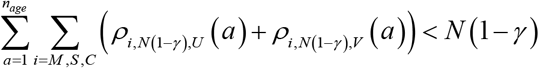, the probability of a major outbreak lies between 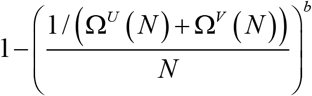 and 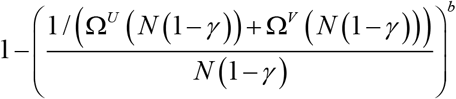.

#### Case 2

For 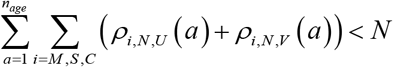 and 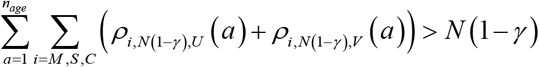, the probability of a major outbreak lies between 0 and 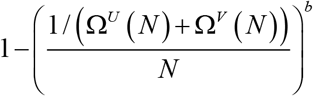.

#### Case 3

For 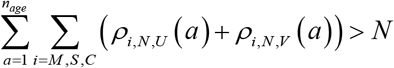, the probability of a major outbreak is 0.

**Figure S2.**
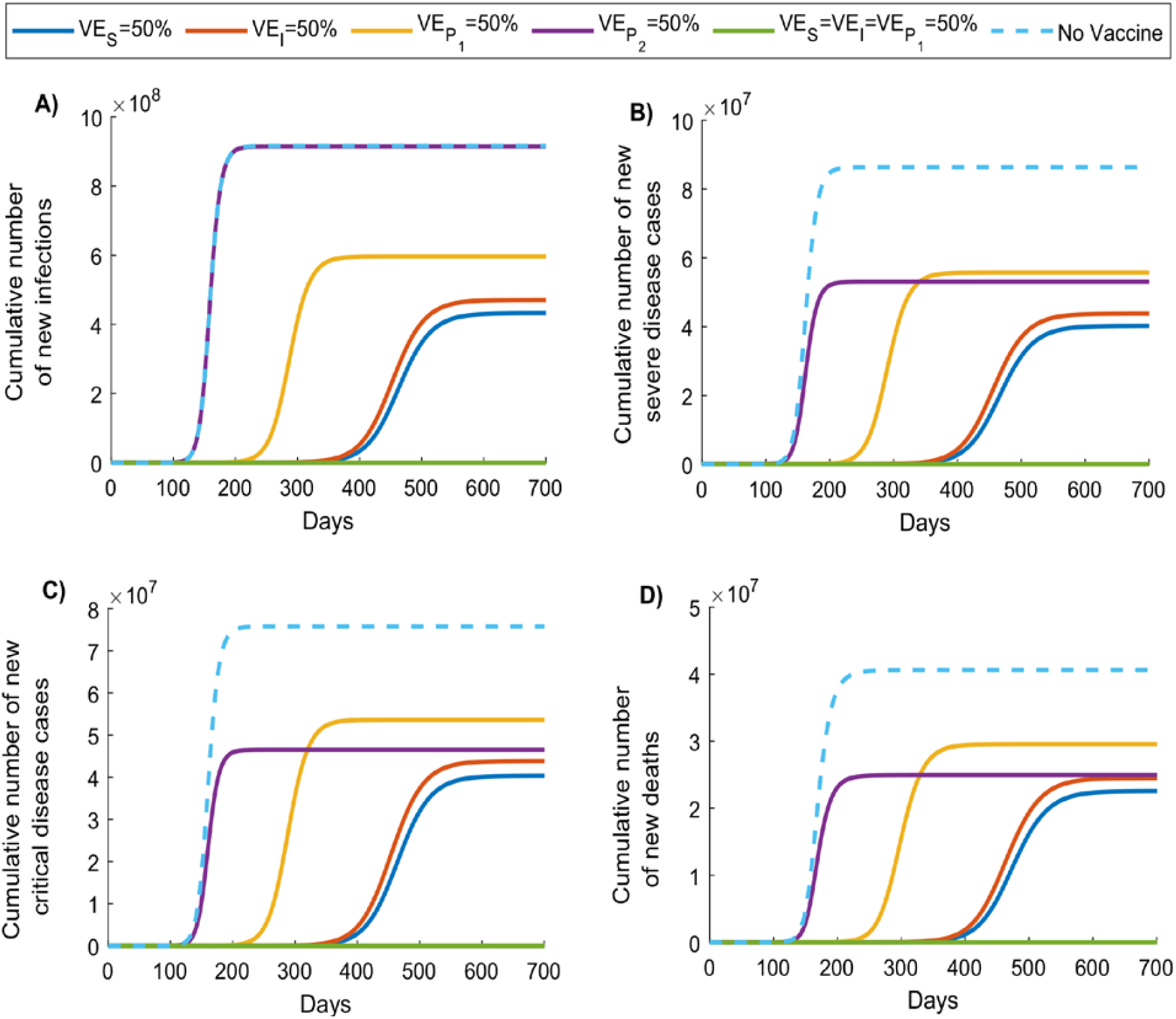
Impact of SARS-CoV-2 vaccination on the cumulative number of A) new infections, B) new severe disease cases, C) new critical disease cases, and D) new deaths in the scenario assuming vaccine scale-up to 80% coverage before epidemic onset. Duration of vaccine protection is 10 years. Impact was assessed at 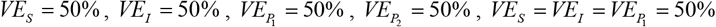.

**Figure S3.**
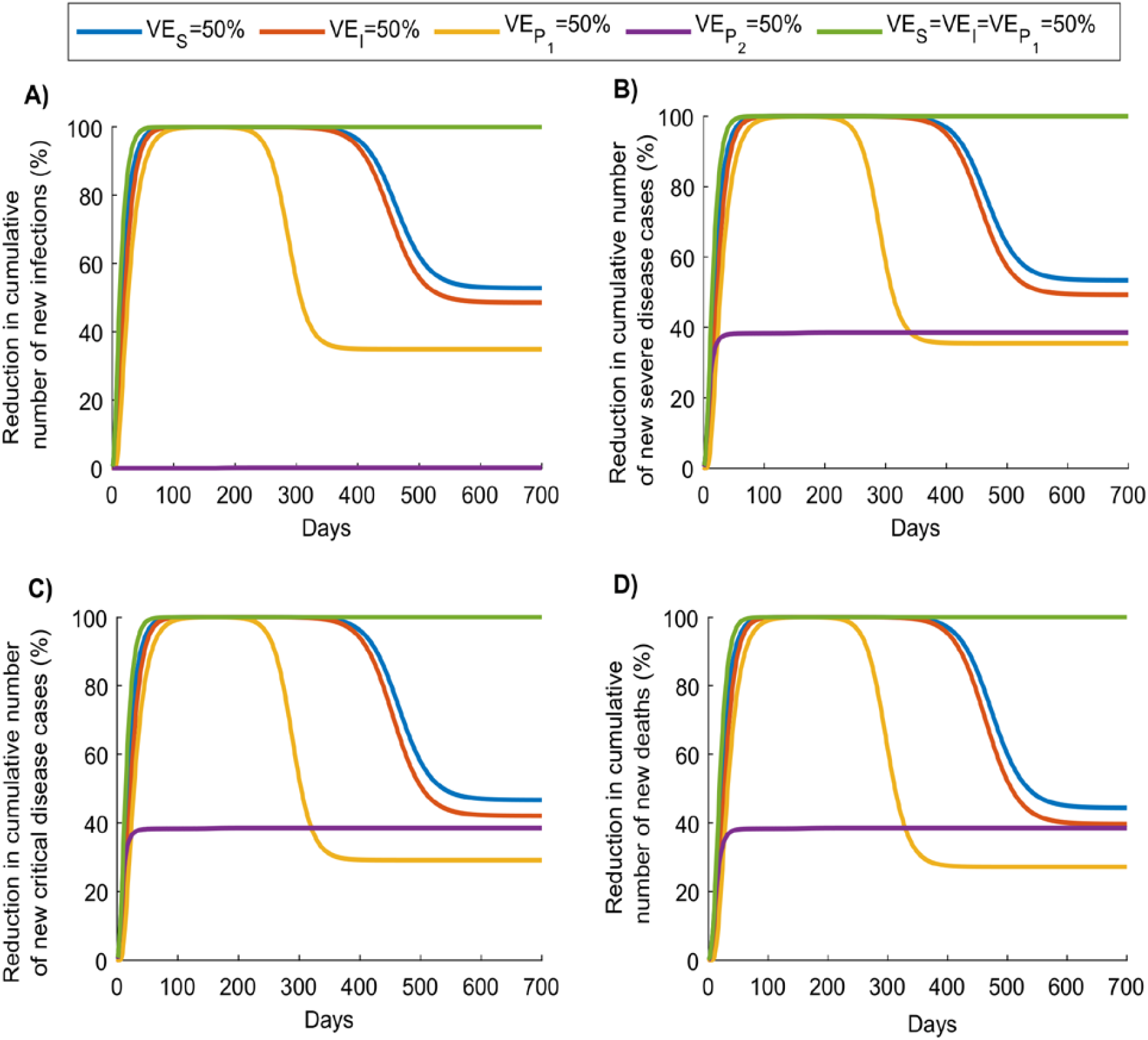
Role of SARS-CoV-2 vaccination in reducing the cumulative number of A) new infections, B) new severe disease cases, C) new critical disease cases, and D) new deaths in the scenario assuming vaccine scale-up to 80% coverage before epidemic onset. Duration of vaccine protection is 10 years. Impact was assessed at 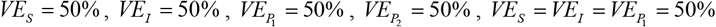.

**Figure S4.**
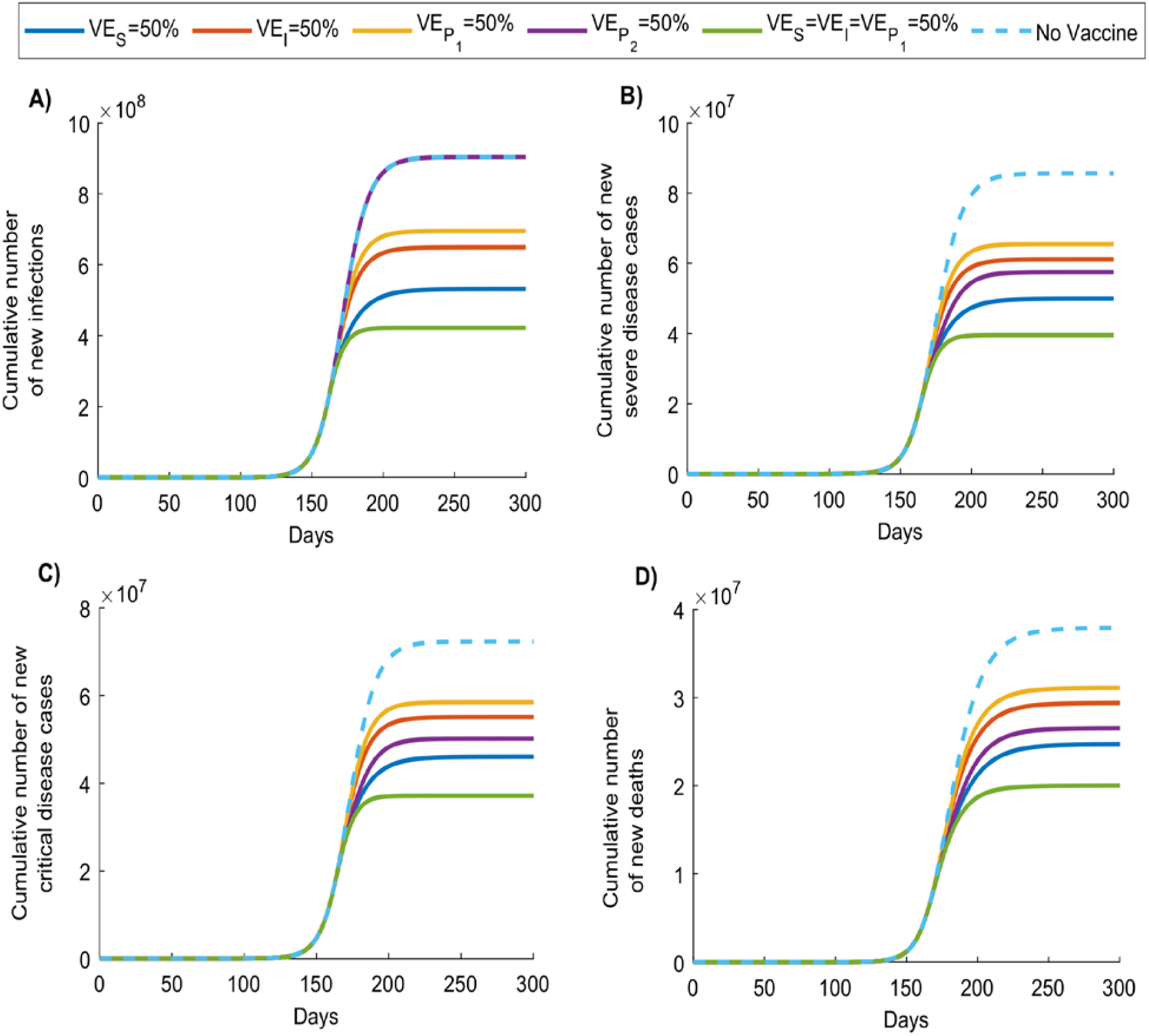
Impact of SARS-CoV-2 vaccination on the cumulative number of A) new infections, B) new severe disease cases, C) new critical disease cases, and D) new deaths in the scenario assuming vaccine introduction during the exponential growth phase of the epidemic, with scale-up to 80% coverage within one month. Duration of vaccine protection is 10 years. Impact was assessed at 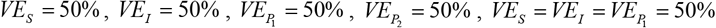.

**Figure S5.**
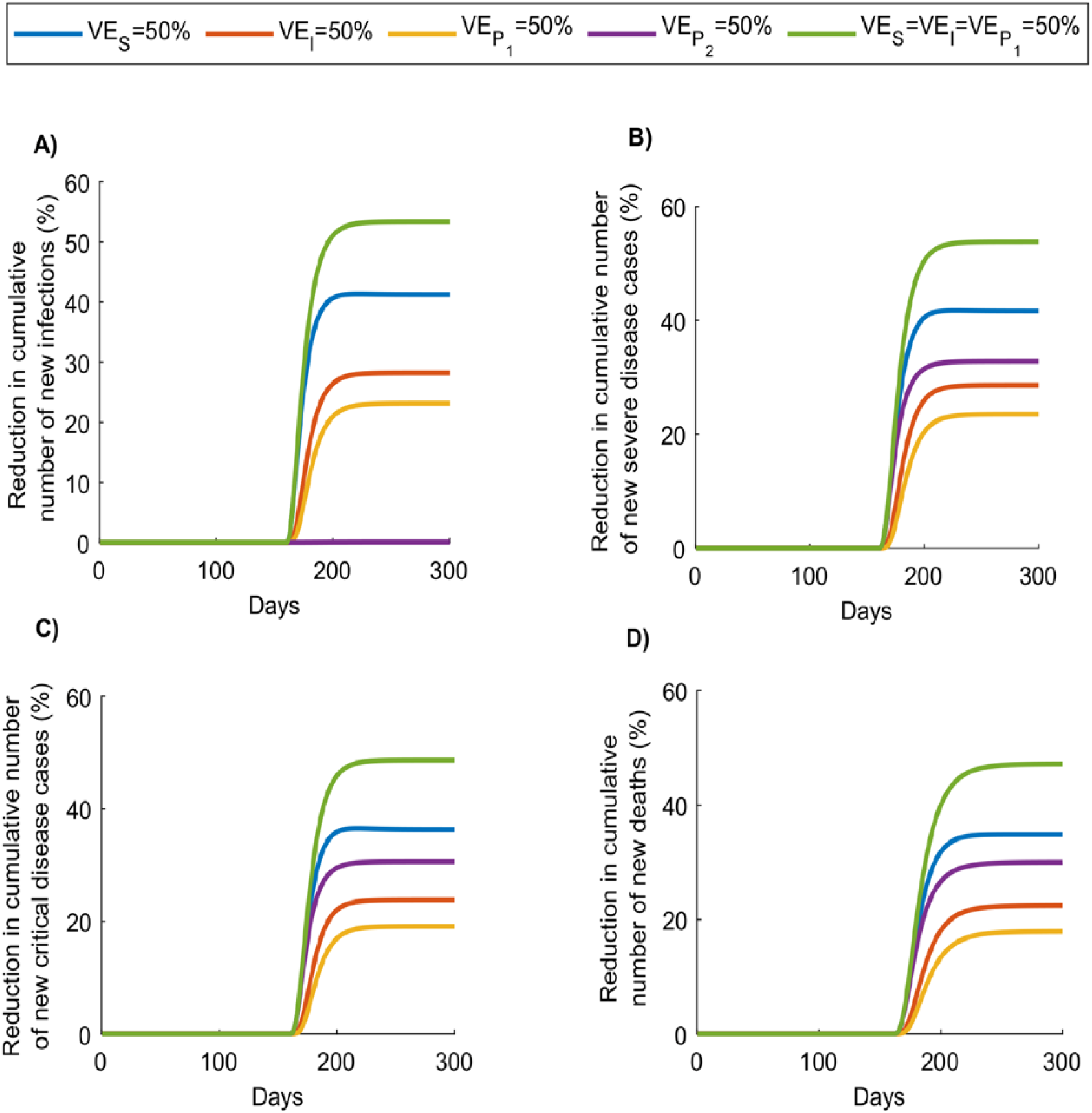
Role of SARS-CoV-2 vaccination in reducing the cumulative number of A) new infections, B) new severe disease cases, C) new critical disease cases, and D) new deaths in the scenario assuming vaccine introduction during the exponential growth phase of the epidemic, with scale-up to 80% coverage within one month. Duration of vaccine protection is 10 years. Impact was assessed at 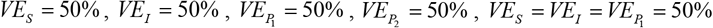.

**Figure S6.**
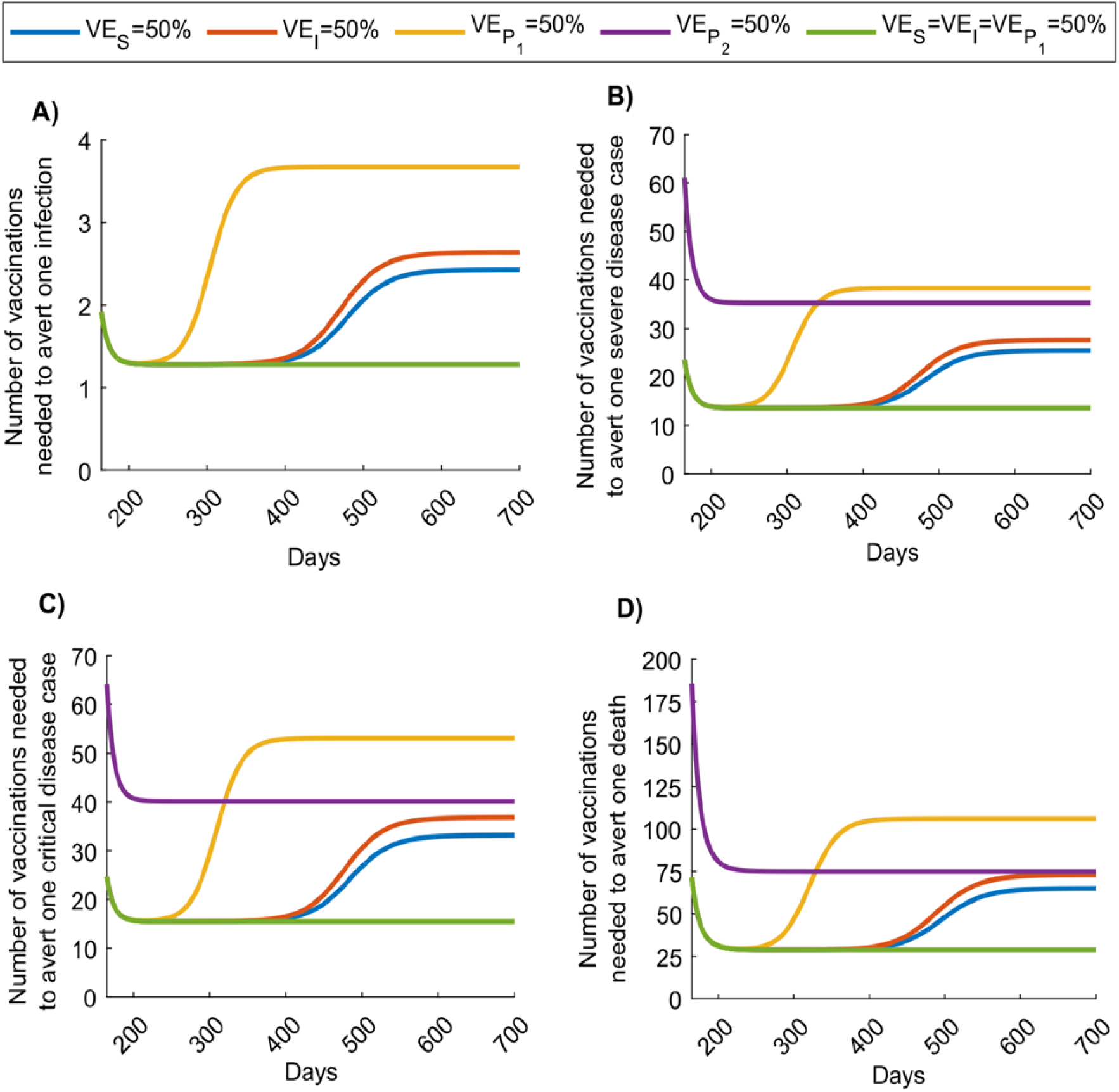
Temporal evolution of SARS-CoV-2 vaccine effectiveness in the scenario assuming vaccine scale-up to 80% coverage before epidemic onset. Number of vaccinations needed to avert A) one new infection, B) one new severe disease case, C) one new critical disease case, and D) one new death, depending on time into the epidemic. Duration of vaccine protection is 10 years. Impact was assessed 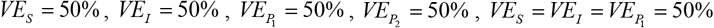. Panel A does not include the result for 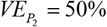, as this efficacy has no impact on number of infections—it affects only severe and critical disease and death.

**Figure S7.**
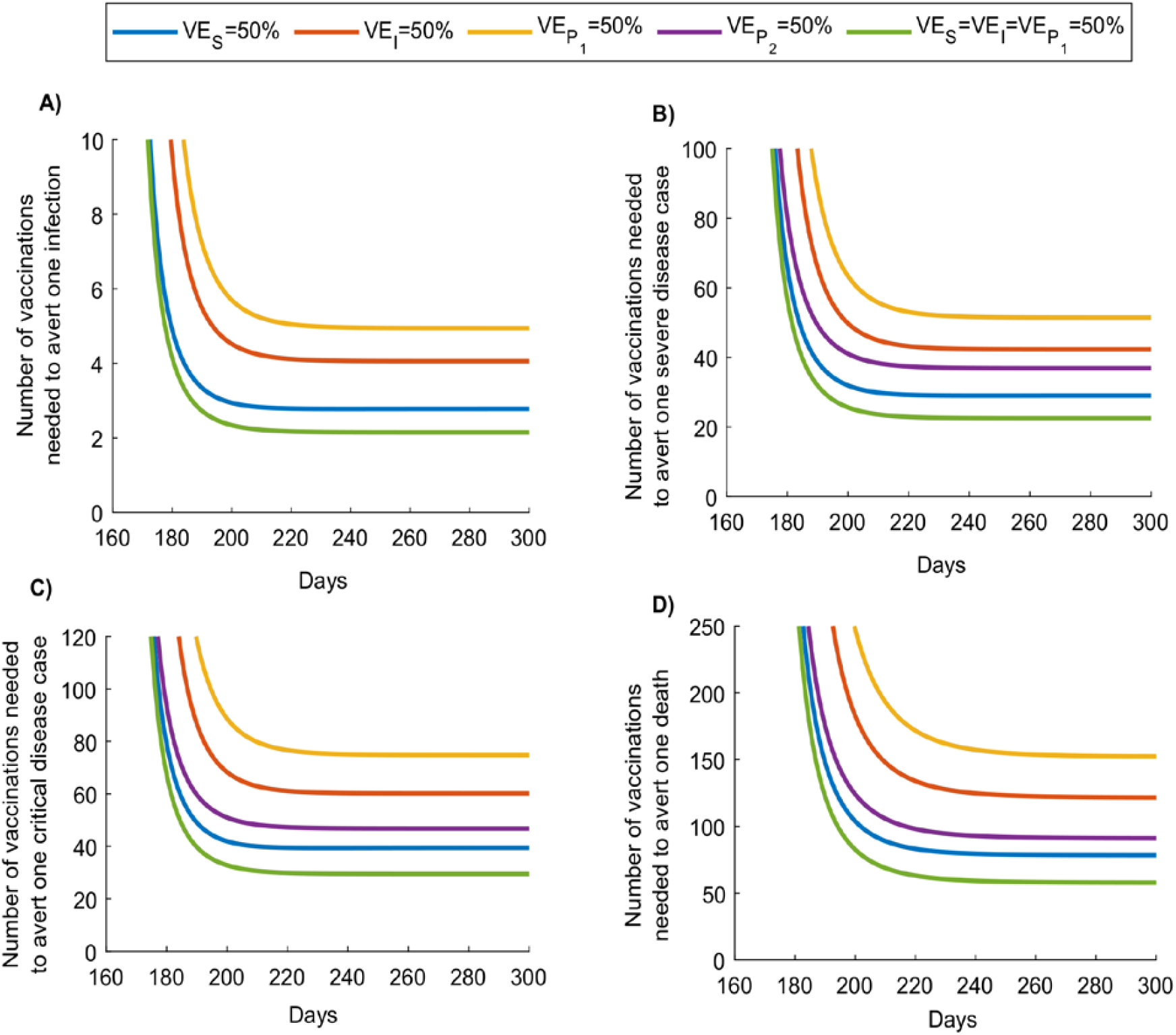
Temporal evolution of SARS-CoV-2 vaccine effectiveness in the scenario assuming vaccine introduction during the exponential growth phase of the epidemic, with scale-up to 80% coverage within one month. Number of vaccinations needed to avert A) one new infection, B) one new severe disease case, C) one new critical disease case, and D) one new death, depending on time into the epidemic. Duration of vaccine protection is 10 years. Impact was assessed at 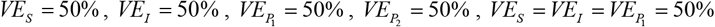. Panel A does not include the result for 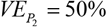, as this efficacy has no impact on number of infections—it affects only severe and critical disease and death.

**Figure S8.**
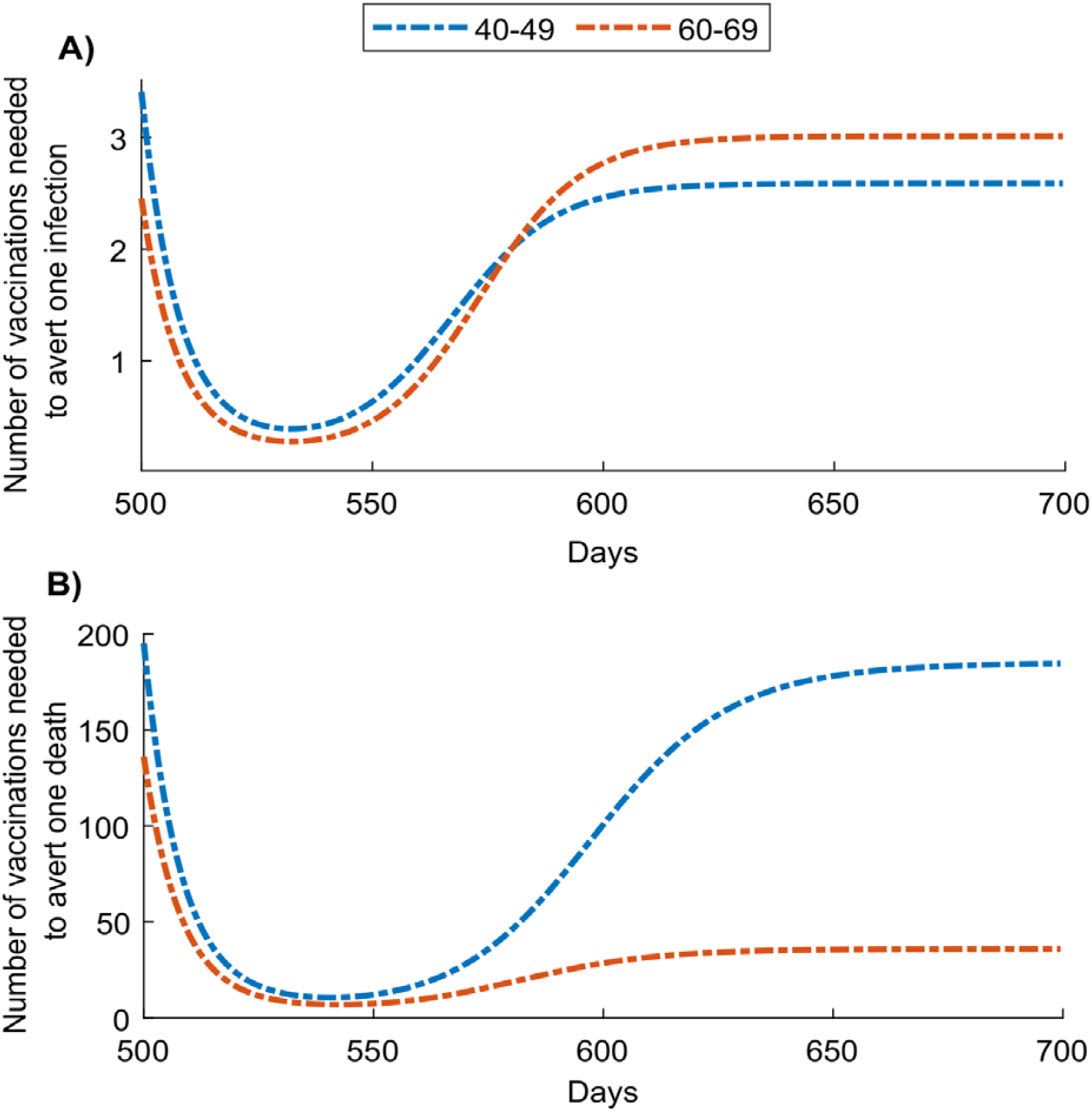
Temporal evolution of effectiveness of age-group prioritization using a SARS-CoV-2 vaccine with *VE*_*s*_ of 50%. Number of vaccinations needed to avert A) one infection and B) one death, by prioritizing individuals aged 40-49 or 60-69 years. Scenario assumes vaccine scale-up to 80% coverage before epidemic onset. Duration of vaccine protection is 10 years.

**Figure S9.**
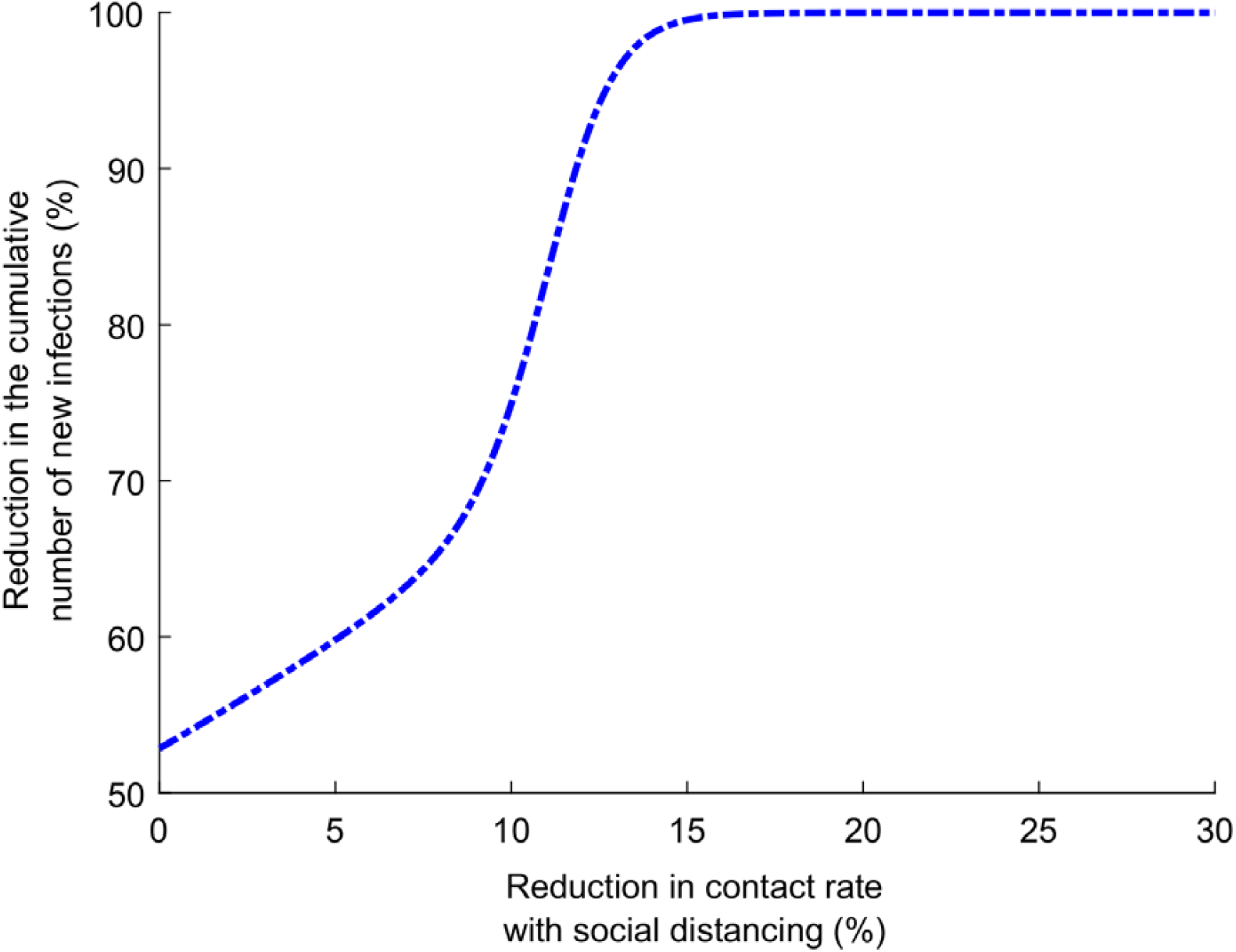
Impact of a social-distancing intervention reducing the contact rate in the population on the cumulative number of new SARS-CoV-2 infections, when introduced to supplement the impact of a vaccine that has 50% efficacy in reducing susceptibility, *VE*_*S*_. Scenario assumes vaccine scale-up to 80% coverage before epidemic onset. Duration of vaccine protection is 10 years.

**Figure S10.**
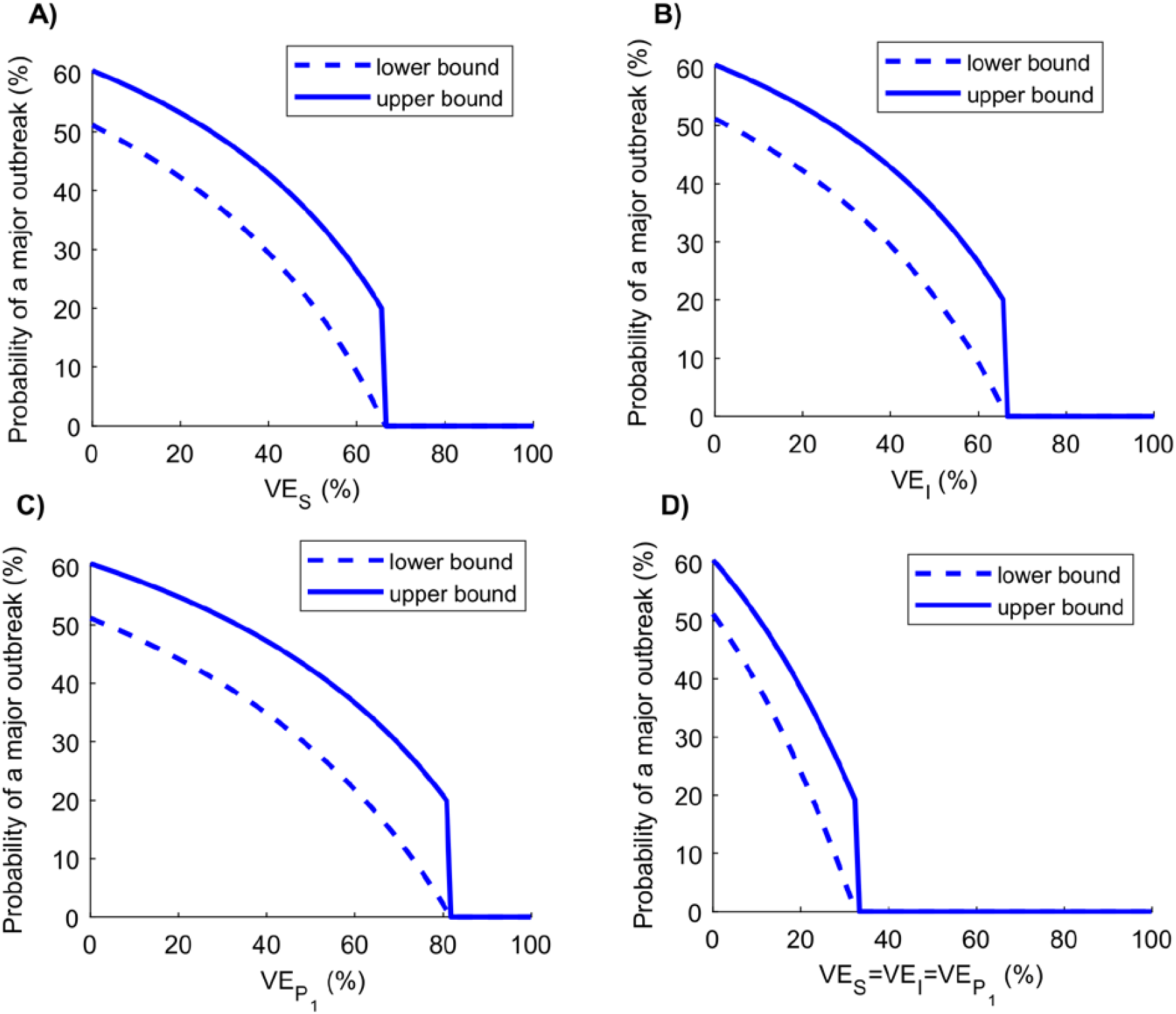
Probability of occurrence of a major outbreak following vaccination. Upper (solid line) and lower (dashed line) bounds of the probability of occurrence of a major outbreak upon virus introduction at varying levels of A) *VE_S_*, B) *VE_I_*, C)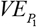, and D) 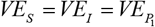. Scenario assumes vaccine scale-up to 80% coverage before epidemic onset. Duration of vaccine protection is 10 years. Figure does not include the result for 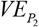, as this efficacy has no impact on probability of occurrence of a major outbreak.

**Figure S11.**
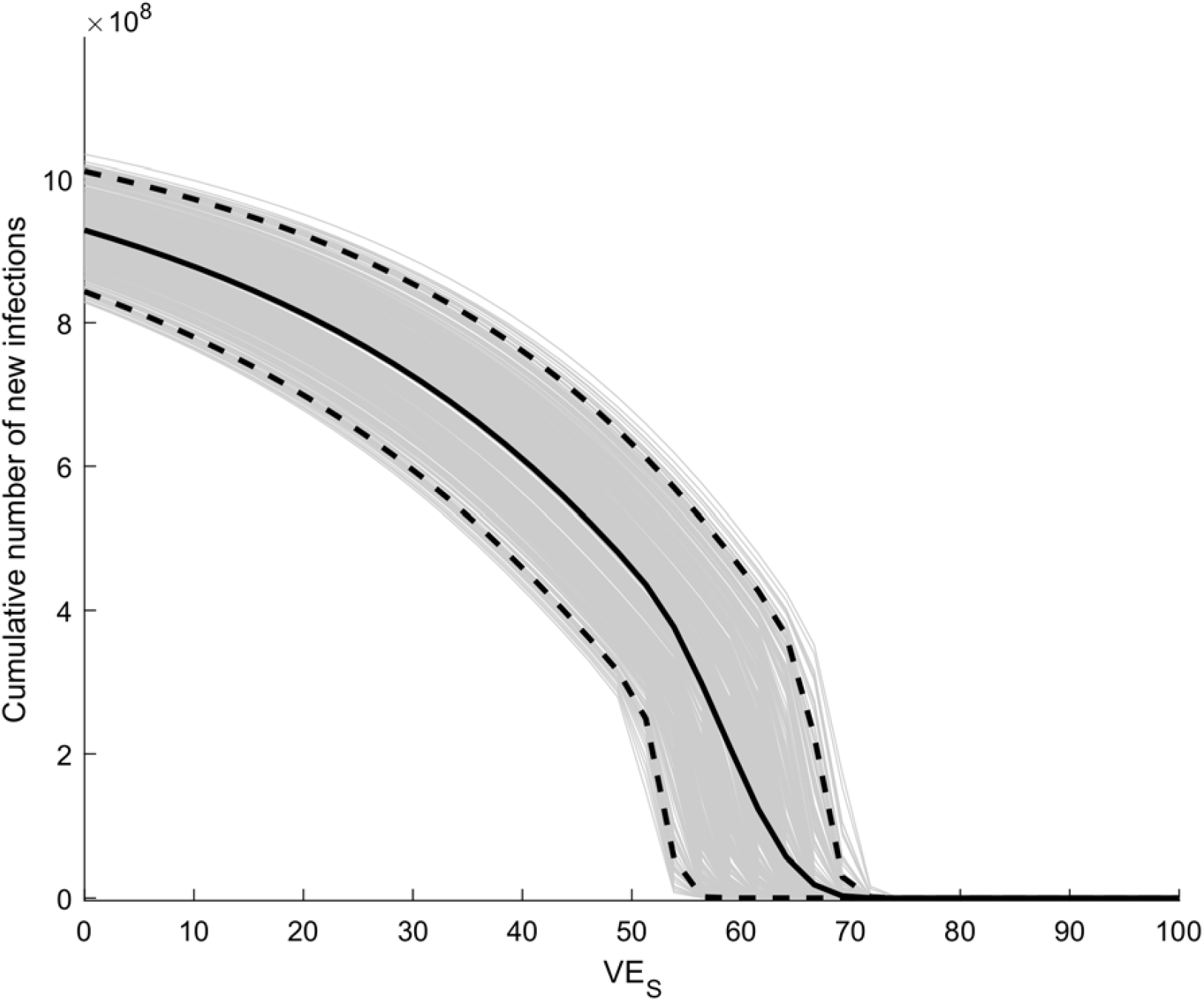
Uncertainty analysis. Model predictions for the mean cumulative number of new infections and associated 95% uncertainty interval (UI) at various levels of vaccine efficacy in reducing susceptibility (*VE*_*S*_) generated through 500 simulation runs. Scenario assumes vaccine scale-up to 80% coverage before epidemic onset. Duration of vaccine protection is 10 years. The solid black line, dashed lines, and shades show respectively, the mean, 95% uncertainty interval, and individual estimates for the cumulative number of new infections across all 500 uncertainty runs.

**Figure S12.**
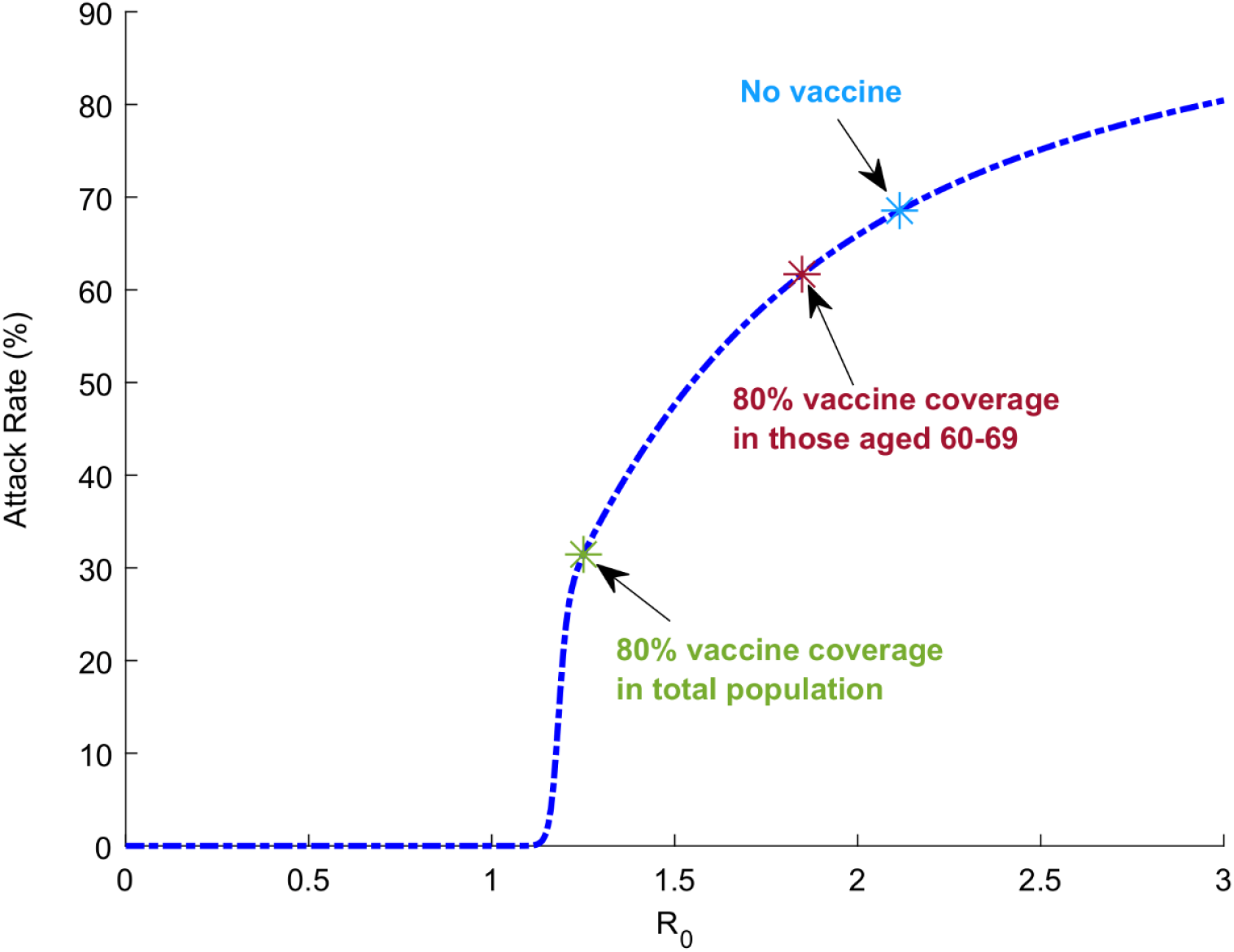
Vaccine effectiveness of age-group prioritization and the reproduction number *R*_0_. Model predictions for SARS CoV-2 attack rate at various levels of *R*_0_, indicating also the impact on *R*_0_ of prioritizing those 60-69 years of age for vaccination or vaccinating all age groups. The blue dashed-dotted line shows the model-predicted attack rate at various levels of *R*_0_. The blue, red, and green stars show, respectively, the model-predicted attack rates in absence of vaccination, by prioritizing vaccination at 80% coverage for those 60-69 years of age, and by extending vaccination at 80% coverage to all age groups. The figure highlights how effectiveness of the vaccine (number of vaccinations needed to avert one infection) depends on the position on the *R*_0_ curve—prioritizing vaccination for any single age group, regardless of that age group, has overall lower effectiveness than extending vaccination to all age groups. The reason is that vaccinating one age group reduces *R*_0_ only marginally, whereas vaccinating all age groups reduces *R*_0_ to an epidemic domain where small reductions in *R*_0_ can have more substantial impact on epidemic size.

**Figure S13.**
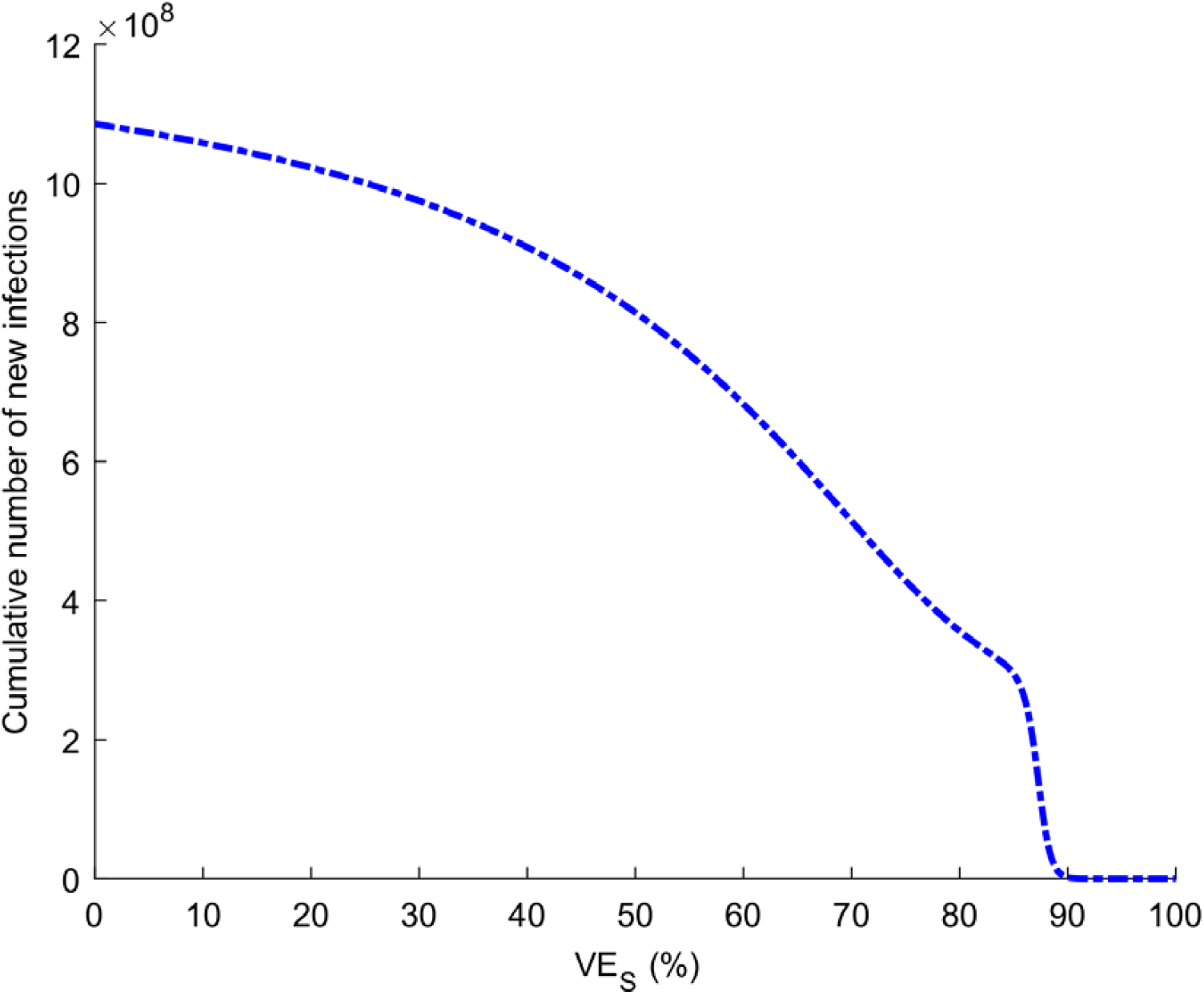
Impact of varying levels of vaccine efficacy in reducing susceptibility, *VE*_*S*_, on the cumulative number of new SARS-CoV-2 infections when the reproduction number *R*_0_ is 3. Scenario assumes vaccine scale-up to 80% coverage before epidemic onset. Duration of vaccine protection is 10 years.

